# Worldwide Prevalence and Clinical Characteristics of RAS Mutations in Head and Neck Cancer: A Systematic Review and Meta-Analysis

**DOI:** 10.1101/2021.11.30.21267087

**Authors:** Ofra Novoplansky, Sankar Jagadeeshan, Ohad Regev, Idan Menashe, Moshe Elkabets

## Abstract

**Importance:** There is considerable variation among different studies for the prevalence of RAS mutations in head and neck cancer (HNC) patients. In light of the development of RAS inhibitors, a reliable assessment of the prevalence of RAS mutations and their correlation with the clinical features of patients with HNC is crucially needed.

**Objective:** To assess the worldwide prevalence of HRAS, KRAS, and NRAS mutations in HNC in the relation to geographical region, anatomical site(s) of the tumor(s) and clinical features.

**Data Sources:** A systematic search of the PubMed, Embase, Web of Science, and Cochrane Central Register of Controlled Trials databases was performed to identify studies published since January 2000. Data were analyzed between June and September 2021.

**Study Selection:** Studies that included mutational analyses of at least one of the target genes and reported the prevalence and frequency of mutations as an outcome measure were included. Studies including less than ten patients or were conducted before year 2000 were excluded.

**Data Extraction and Synthesis:** Two researchers independently reviewed the literature according to Preferred Reporting Items for Systematic Reviews and Meta-analyses (PRISMA) reporting guidelines. Random-effects models were applied to results with high heterogeneity. Otherwise, fixed-effects models were used for the analyses. Dichotomous variables were pooled as odds ratios (OR).

**Main Outcome(s) and Measure(s):** The primary outcome was mutation prevalence. Secondary outcomes included the location of the mutated codon and amino acid substitution.

**Results:** The estimated mutation rate is highest for HRAS (7%), followed by KRAS (2.89%) and NRAS (2.20%). HRAS prevalence in South Asia (15.28%) is twice as high as the global estimate. HRAS mutations are more prevalent in oral cavity and salivary gland tumors. In contrast, KRAS mutations are found more frequently in sinonasal tumors, and NRAS mutations are found chiefly in tumors of the nasopharynx. OR analyses show a significant association between HRAS mutations and a high tumor stage (OR=3.63). In addition, there is a significant association between HPV-positive status and KRAS mutations (OR=2.09).

**Conclusions and Relevance:** RAS mutations occur in a subset of HNC patients and their prevalence varies according to geography, tumor’s anatomical site, stage, and HPV status. This meta-analysis provides support for their potential as viable therapeutic targets in HNC patients.

**Key Points:** *Question:* What is the prevalence of mutations in RAS genes in head and neck cancer (HNC) with respect to anatomical site, geographical region, and clinical features?

*Findings:* This meta-analysis compiles the findings of 149 studies with over 8500 HNC patients and assesses the global prevalence of mutations in the HRAS, KRAS and NRAS genes. The prevalence of RAS mutations varies between the seven major anatomical sites of HNC. The HRAS mutation rate in South Asia is double that in other geographical regions. Mutations in HRAS are associated with advanced disease, and mutations in KRAS are positively associated with HPV status.

*Meaning:* This study presents the most comprehensive assessment of the prevalence of RAS mutations in HNC and their correlation with geographical regions and with clinical features.

## Introduction

The heterogeneous nature of head and neck cancer (HNC)^1^ may explain the limited efficiency of current systemic therapies. There is thus a pressing need to study specific and less common genetic alterations that may affect disease characteristics and clinical outcomes in HNC patients. Proteins of the RAS GTPase family proteins control cell proliferation, differentiation, and survival,^2^ with Kirsten rat sarcoma viral oncogene homolog (KRAS), Neuroblastoma Rat Sarcoma Virus (NRAS), and Harvey rat sarcoma viral oncogene homolog (HRAS) sharing significant sequence homology and largely overlapping functions.^3^ Gain-of-function mutations in RAS genes are found in ∼19% of human cancers,^4^ prompting interest in identifying anti-RAS therapeutic strategies for cancer treatment. The immense effort that has been invested in developing RAS inhibitors has led to several breakthroughs in recent years, allowing for targeted treatment of patients with alterations in RAS genes.^5,6^ Mutations in members of the RAS gene family are seemingly rare in some large cohorts of HNC patients, but a broader examination of all the available studies reveals large variations in the assessment of the prevalence of the different mutations. Here, we present a systematic review and meta-analysis of the prevalence of RAS mutations in HNC patients in the contexts of geographical region, tumor anatomical site and clinical features. These findings highlight RAS as a potential therapeutic target in some HNC patients.

## Methods

This systematic review adhered to the Preferred Reporting Items for Systematic Reviews and Meta-Analyses (PRISMA) Checklist.^7^

### Study design

We evaluated the prevalence of mutations in HRAS, KRAS, and NRAS genes in HNC patients.

### Search Strategy

A systematic review of the literature was conducted by searching the PubMed, Embase, Web of Science, and Cochrane Central Register of Controlled Trials databases in June 2021 for studies published in the English language since 1 January 2000. The strings used are in the systemic search of databases are detailed in eMethods in the Supplement. The bibliographies of retrieved studies and systematic reviews identified in the search were screened for relevant references. Publicly available databases were screened for unpublished data.

### Selection Criteria

The inclusion criteria for the meta-analysis were that the study had to include a mutational analysis of at least one of the target genes (HRAS, KRAS, or NRAS) and a report the prevalence and frequency of mutations as an outcome measure. Exclusion criteria were defined as: (1) Studies displaying results from patients with tumors other than HNCs or mutations other than those in the target genes; (2) Studies that did not report data related to the prevalence or frequency of mutations in the target genes; (3) Studies that did not evaluate target genes for somatic mutations; (4) Studies published before 1 January 2000; (5) Studies that were conducted using cell lines or animal models; (6) Studies of pediatric populations; (7) Review articles, letters, personal opinions, book chapters, or conference abstracts; (8) Studies containing data included in other studies or studies in which it was not possible to determine whether duplicate data were included; and (9) Studies enrolling fewer than ten patients.

### Data Extraction

Two researchers (SJ, ON) screened the studies at the title and abstract level, followed by a full-text review. Disagreements over inclusion were resolved by consensus adjudication, and studies were extracted into a standardized extraction database. Extracted variables included study cohort size, number of mutated cases for each RAS family gene, primary tumor location, tumor grade or stage, geographical origin of studied patients, mutation assessment method, mutated codon, HPV-status, and biopsy type, if reported.

### Evaluation of Quality and Risk of Bias

Our study selection process excluded individual case reports and cohorts of less than ten patients due to the risk of bias. All papers considered after initial screening were reviewed and scored for risk of bias according to the Joanna Briggs Institute Critical Appraisal Checklist for Studies Reporting Prevalence Data^8^. Studies that did not evaluate all three RAS family members were considered more prone to risk of bias and were not included in the general prevalence analysis. In addition, publication bias and heterogeneity were assessed by visual inspection of funnel plots and via Egger’s regression test^9^ (eFigure 1 in the Supplement).

**Figure 1.**
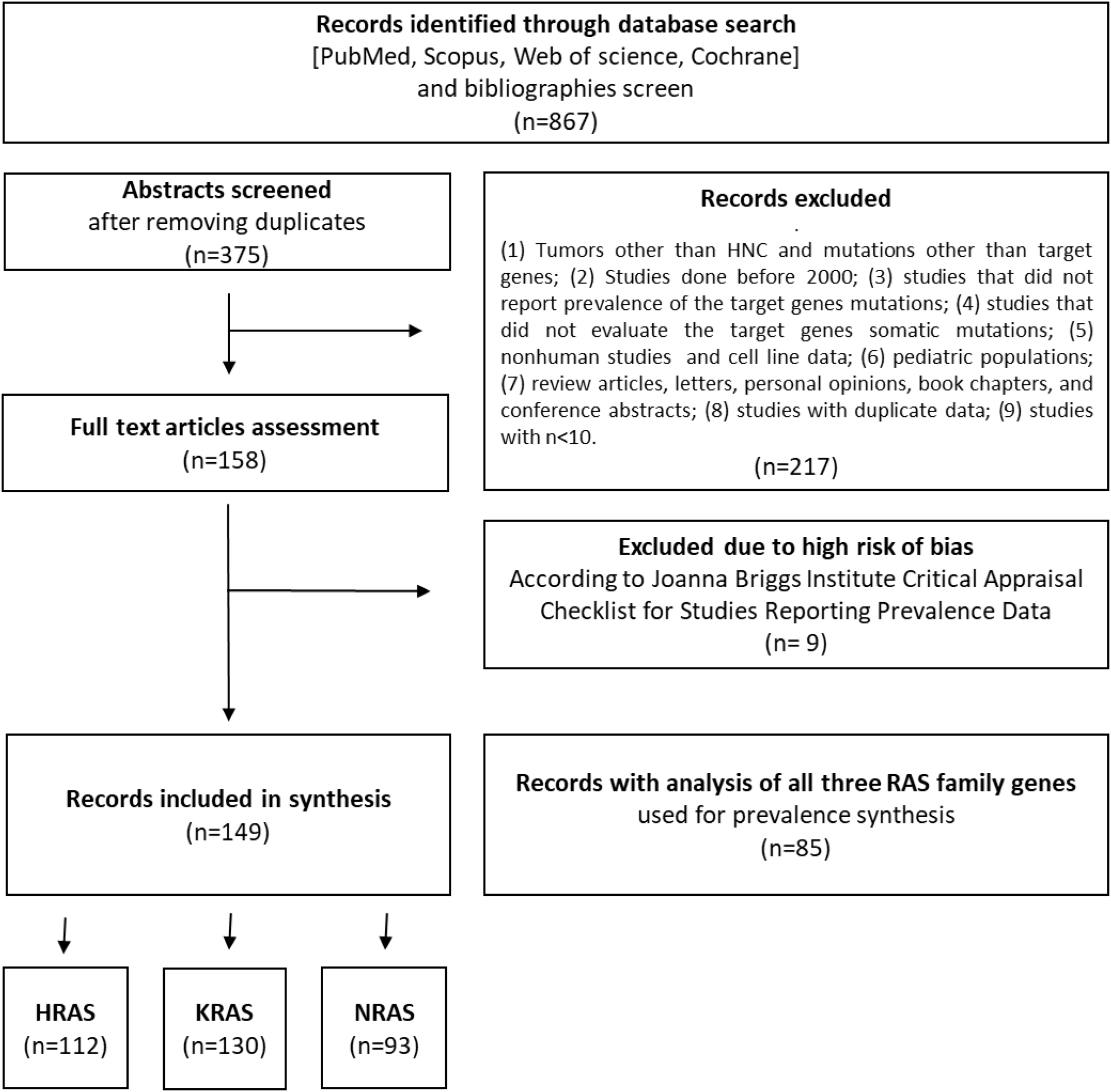
Flow diagram of the literature search process and selection criteria.

### Statistical Analysis

Pooled prevalence rates, pooled odds ratios (ORs), and forest plots were generated using the R Meta and MetaFor Packages.^10,11^ The Cochrane Q chi-squared test and the inconsistency index statistic (I^2^) were used to examine the heterogeneity across studies. Fixed-effects models were used to assess the pooled prevalence of genes for results with low heterogeneity (I^2^ ≤ 50%). Otherwise, random-effects models were used for the analyses. A sensitivity analysis using a “leave-one-out” paradigm from the built-in function in MetaFor, as proposed by Wang et al.,^12^ was used to assess each study’s effect on the overall pooled prevalence and detected outliers.^12^ First, the pooled overall prevalence of mutations in the three different target genes (KRAS, HRAS, NRAS) was calculated with a corresponding 95% confidence interval (95% CI). Next, subgroup analyses were performed according to geographical region and anatomical site. Finally, we assessed the association between the RAS gene mutational status and HPV status or tumor grade using the R MetaBin function.

## Results

### Study Selection

The flow diagram shown in **Figure 1** depicts the search strategy and study selection process. A total of 867 studies were retrieved from four electronic databases and a bibliography screen. After the removal of duplicates, 375 studies were considered potentially eligible for evaluation, but 217 did not meet all the inclusion criteria, leaving a final sample of 158 studies. Nine additional studies were excluded due to the high risk of bias. To reduce the risk of bias, only papers with the highest grade (n = 85) were included for pooled analyses of the overall mutation prevalence. The literature references for the studies included in the meta-analysis are listed in eTable 1 in the Supplement.

### Study Characteristics

Detailed characteristics of the studies are provided in eTable 2 in the Supplement. Of the 149 studies included in the analysis, 112, 130, and 93 contained data pertaining to gene alterations in HRAS, KRAS, and NRAS, respectively, and 85 presented analyses of all three RAS family genes. In total, 148 of the included studies were cohort studies, while one was a phase 1 clinical trial. Forty-seven studies used targeted next-generation sequencing, 46 utilized Sanger sequencing, 23 employed whole-exome sequencing, 9 conducted Mass Array analysis, 4 used whole-genome analyses, and 20 employed other or mixed analysis methods. The anatomical location of the tumors in the included study cohorts are detailed in eTable 3 in the Supplement. The studies were conducted in 29 different countries. Four studies included mixed populations from various geographical regions.

### Risk of Bias within Studies

Nine studies were classified as having a high risk of bias and were therefore excluded from this meta-analysis. Eleven studies were classified as having a moderate risk of bias due to a small cohort size, while 57 studies were classified as having a moderate risk of bias since they analyzed only one of the three RAS family target genes. The remaining 85 studies were classified as having a low risk of bias and were used in the general prevalence analysis. All low and moderate risk studies were used in prevalence analyses pertaining to tumor anatomical sites, mutated codons, and the association between RAS mutations and patient clinical features.

### Prevalence of RAS Mutations

#### HRAS mutations

Mutations in HRAS were identified in 564 tumors from 8501 patients. The mean prevalence of HRAS mutations was 7% (95% CI, 5.38-9.06, p <0.01, I^2^ = 87%) (**Figure 2A**). Geographical region-specific analyses revealed significant differences in these rates in different regions of the world (Q = 22.51, Pv <0.0001). The mean frequency of HRAS mutations in South Asia was 15.28%, with this rate being higher than the rates in other geographical regions, including East Asia (5.07%), Europe (4.65%), and North America (6.87) (**Figure 3A**, eFigure 2 in the Supplement).

**Figure 2.**
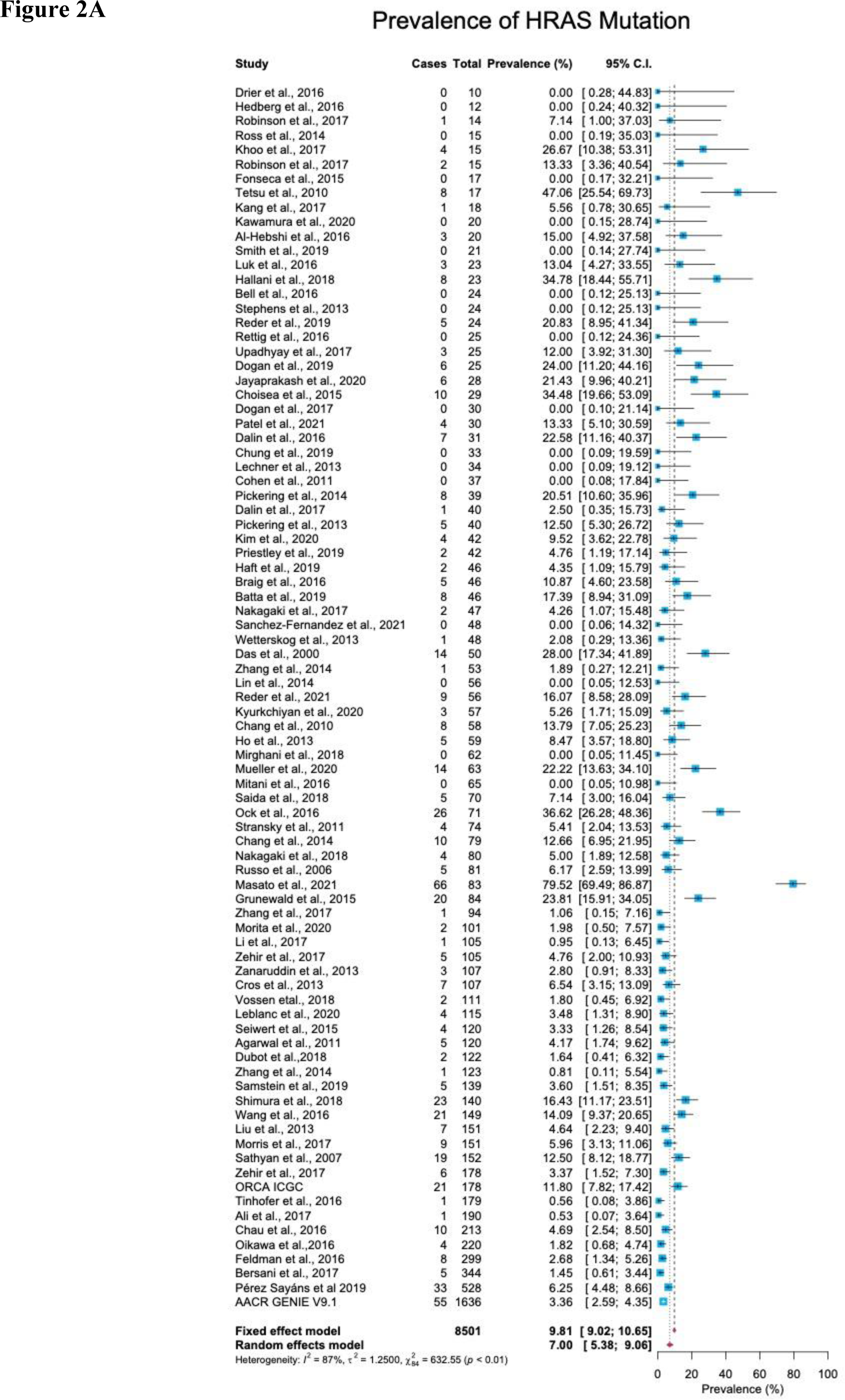
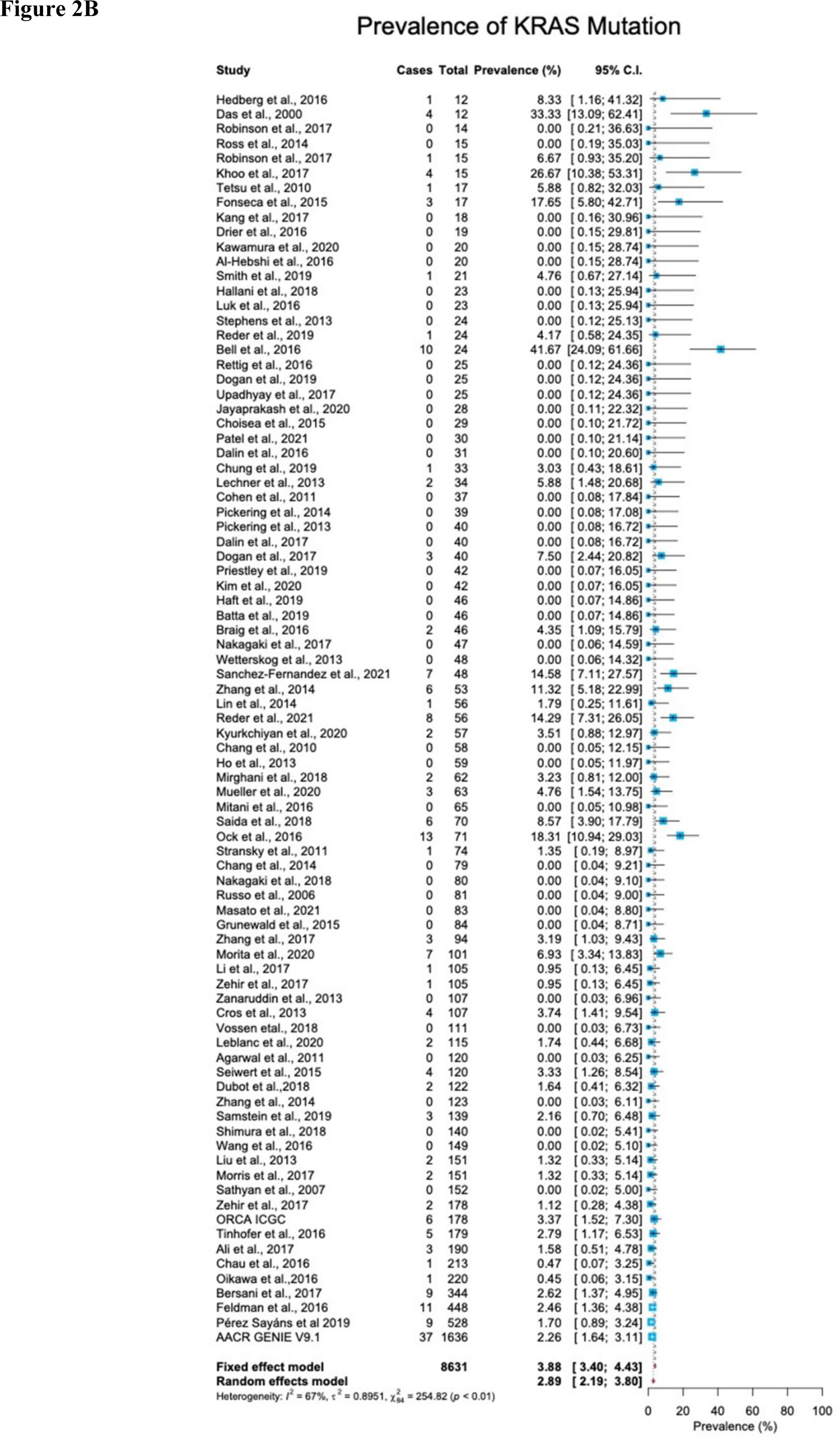
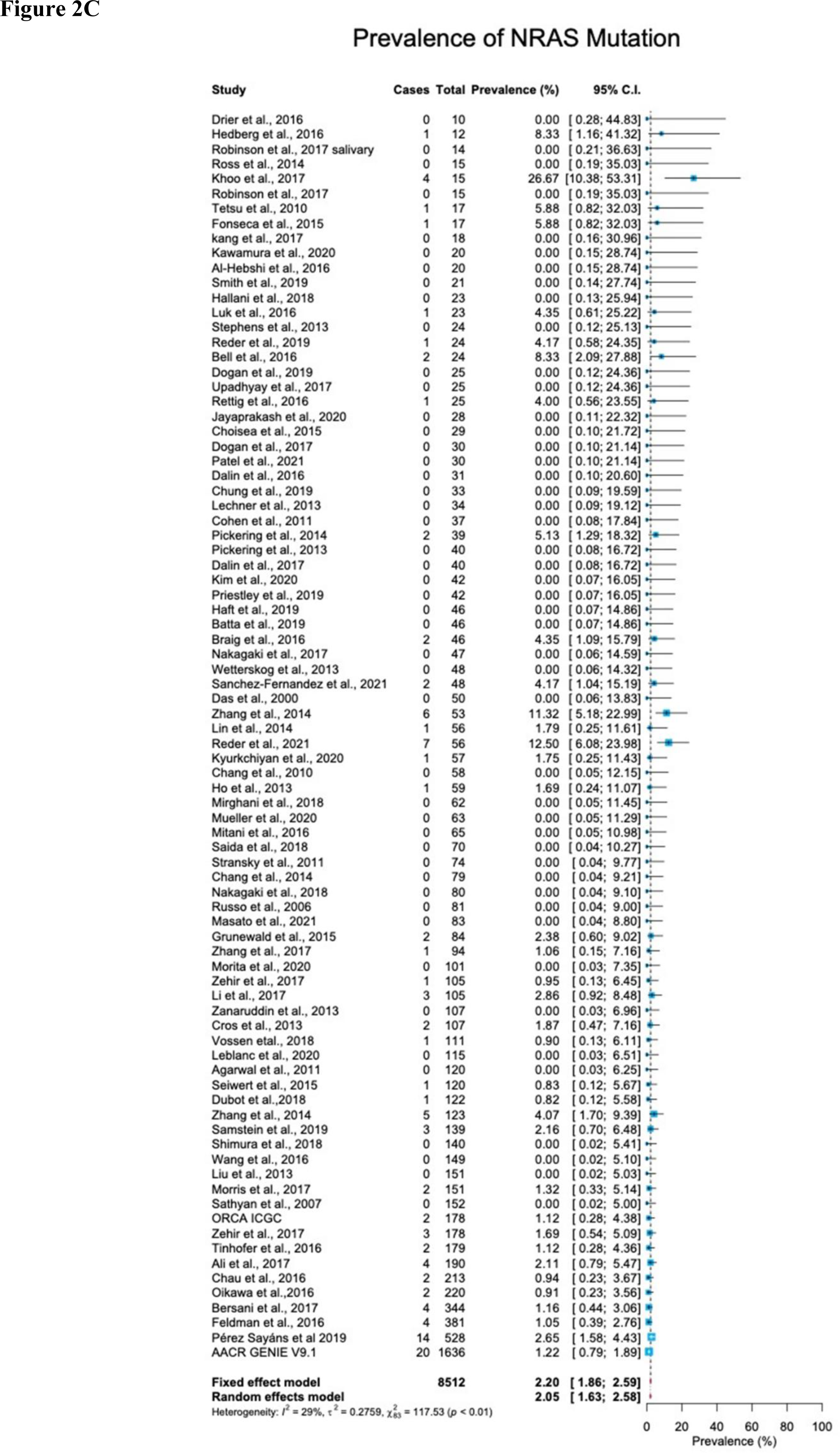
Prevalence of RAS mutations in head and neck cancer. **(A)** The prevalence of HRAS mutations is 7% (95 % CI, 5.38-9.06, p <0.01, I^2^ = 87%). **(B)** The prevalence of KRAS mutations is 2.89% (95 % CI, 2.19-3.80, p <0.0.1, I^2^ = 67%). **(C)** The prevalence of NRAS mutations is 2.20% (95 % CI, 1.86-2.59, p <0.01, I^2^ = 29%). CI: Confidence interval. I^2^: Inconsistency index.

**Figure 3.**
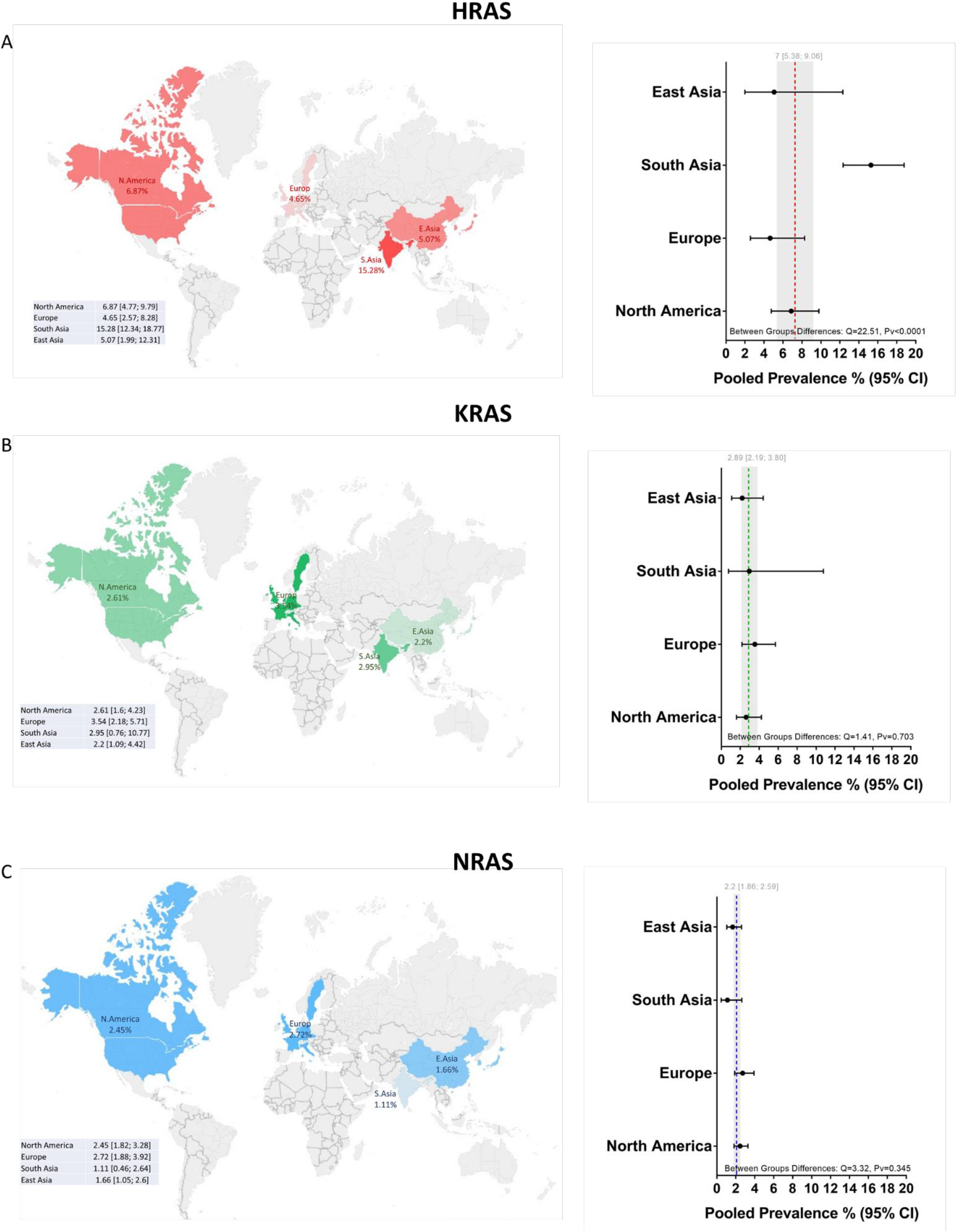
Global prevalence of RAS mutations in head and neck cancer. Cohort studies were grouped according to the geographical origins of the patients. On the map of the world are shown the frequencies [%] of HRAS, KRAS, and NRAS mutations in East Asia, South Asia, Europe, and North America. Dotted lines and gray shading correspond to the overall prevalence and the 95% CI. CI: Confidence interval. I^2^: Inconsistency index.

#### KRAS mutations

Mutations in KRAS were identified in 188 tumors from 8631 patients. The mean prevalence of KRAS mutations was 2.89% (95% CI, 2.19-3.80, p <0.0.1, I^2^ = 67%) (Figure 2B), with no significant differences in prevalence between analyzed geographical regions (Q = 1.41, Pv = 0.7). (Figure 3B, eFigure 2 in the Supplement).

#### NRAS mutations

Mutations in NRAS were identified in 113 tumors from 8512 patients. The mean prevalence of NRAS mutations was 2.20% (95% CI, 1.86-2.59, p <0.01, I^2^ = 29%) (Figure 2C). No significant differences in these rates were observed between the different parts of the world (Q = 3.32, Pv = 0.34) (eFigure 2 in the Supplement).

### Anatomical Site

As HNC includes tumors that arise from a wide range of anatomical sites and sub-sites, an analysis of the frequency of mutations in the three RAS genes was performed for seven major anatomical areas. A summary of these analyses is presented in **Figure 4A** and eFigure 3 in the Supplement.

**Figure 4:**
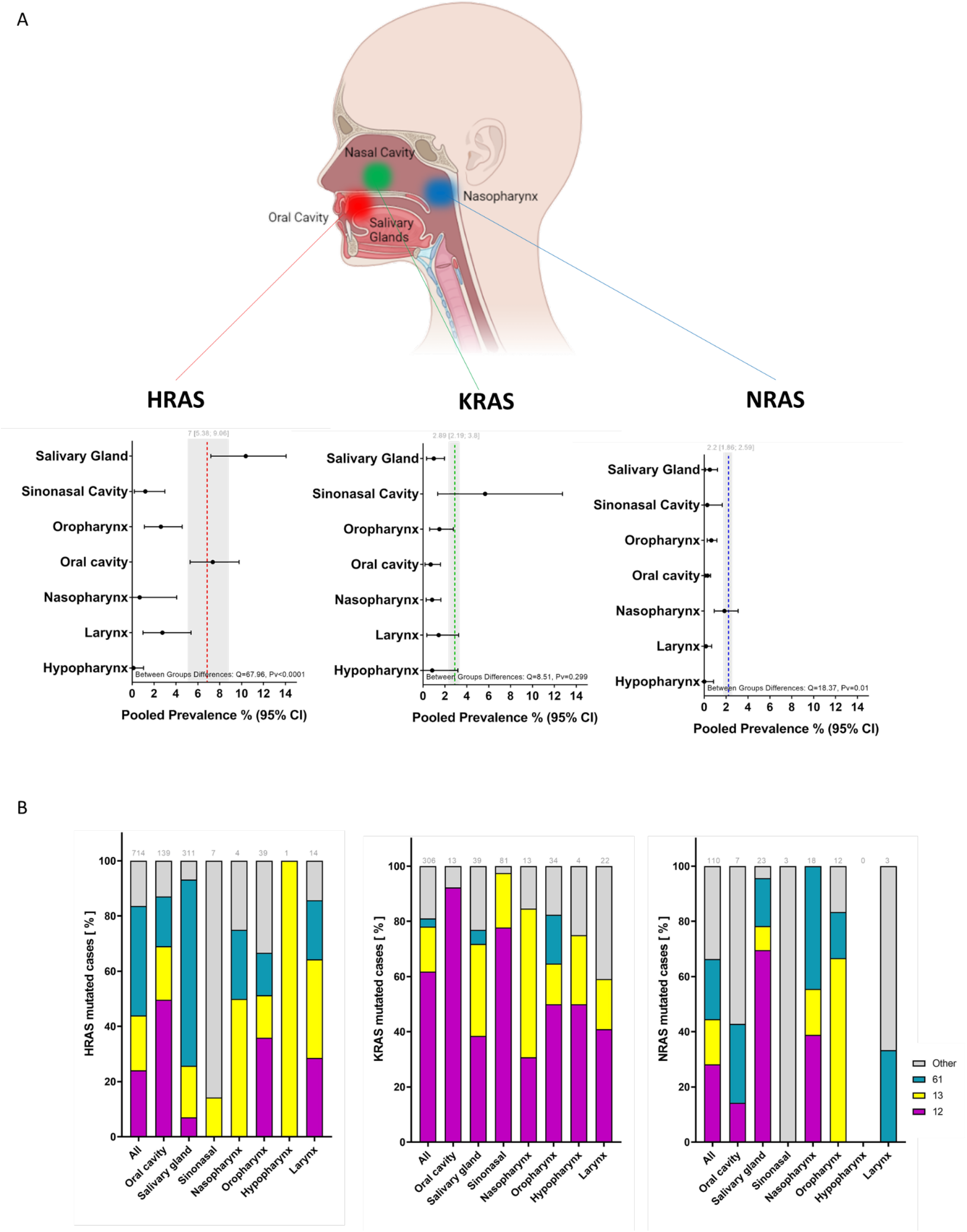
Prevalence of mutations and locations of mutated codons according to tumor anatomical site. **(A)** Prevalence of RAS mutations according to tumor anatomical site. Dotted lines and gray shading correspond to the overall prevalence and the 95% CI. **(B)** Mutated codon locations [%] according to tumor anatomical site. CI: Confidence interval. I^2^: Inconsistency index.

#### HRAS mutations

A significant difference in the prevalence of HRAS mutations was detected between anatomical sites (Q = 67.96, Pv <0.0001): HRAS mutations were found more frequently in tumors of the salivary glands (10.37%; 95% CI, 7.18-14.06) and oral cavity (7.36%; 95% CI, 5.39-9.76) than in tumors of the sinonasal cavity (1.2%; 95% CI, 0.2-3), oropharynx (2.6%; 95% CI, 1.12-4.56), nasopharynx (0.68%; 95% CI, 0-4.06), larynx (2.76%; 95% CI, 0.99-5.38), or hypopharynx (0.12%; 95% CI, 0-0.04).

#### KRAS mutations

A trend towards more frequent KRAS mutations was observed for tumors of the sinonasal cavity (5.67%; 95% CI, 1.33-12.74) as compared to tumors of the salivary glands (0.98%; 95% CI, 0.33-1.96), oral cavity (0.7%; 95% CI, 0.17-1.59), oropharynx (1.49%; 95% CI, 0.6-2.77), nasopharynx (0.83%; 95% CI, 0.29-1.63), larynx (1.43%; 95% CI, 0.34-3.25), or hypopharynx (0.84%; 95% CI, 0-3.18). However, these differences were not robust (Q = 8.5, Pv = 0.29).

#### NRAS mutations

A significant difference between anatomical sites was also seen for NRAS mutations (Q = 18.37, Pv = 0.01), with a rate of 1.85% (95% CI, 0.92-3.1) in the nasopharynx as compared to lower rates in tumors of the salivary glands (0.51%; 95% CI, 0.11-1.22), oral cavity (0.3%; 95% CI, 0.11-0.58), sinonasal cavity (0.28%; 95% CI, 0-1.65), oropharynx (0.65%; 95% CI, 0.28-1.16), larynx (0.16%; 95% CI, 0-0.68), or hypopharynx (0%; 95% CI, 0-0.85).

### Amino Acid Substitution

#### HRAS mutation

In an analysis of all cases with HRAS mutations, 24%, 20%, and 39% were situated in codons 12, 13, and 61, respectively. Salivary gland tumors exhibited a higher frequency of mutations in codon 61 (67%), while in tumors of the oral cavity, mutations in codon 12 were the most frequent (50%) (Figure 4B, left side). Mutations in codon 12 were mostly G12S point mutations (56.3%), while those in codon 13 were primarily G13R point mutations (46.8%). Lastly, mutations found in Q61 were primarily Q61R (49.2%), Q61K (26.4%), and Q61L (22.2%) point mutations (eFigure 4 in the Supplement).

#### KRAS mutations

Mutations in codon 12 were the most frequent across all anatomic sites, followed by codon 13. Mutations in codon 61 were primarily detected in tumors of the oropharynx (17%) (Figure 4B, middle). Among the codon 12 mutations, the most common amino acid substitution was G12D (51%), followed by G12V (16.3%) and G12C (12.9%) (eFigure 4 in the Supplement).

#### NRAS mutation

NRAS mutations were more evenly distributed among the different codons (Figure 4B, right side). We note that site-specific analyses should be interpreted with caution owing to the limited number of mutated cases.

### Association between RAS Mutations and Disease Stage/Grade

Tumor grade and stage are well-studied prognostic factors for HNC.^13^ In total, 44 studies reported details of the tumor stage or grade of patients along with the mutation status. Tumors with a stage or grade of 1 and 2 were defined as low-grade tumors, while those with a stage or grade of 3 and 4 were categorized as high-grade tumors. An OR analysis revealed a significant association between HRAS mutation and advanced stage (OR = 3.63; 95% CI, 1.53-8.64) (**Figure 5A**). KRAS (OR = 2.41; 95% CI, 0.85-6.86) and NRAS (OR = 1.52; 95% CI, 0.68-3.41) mutations were both associated with an OR>1, but did not reach statistical significance (eFigure 5 in the Supplement).

**Figure 5:**
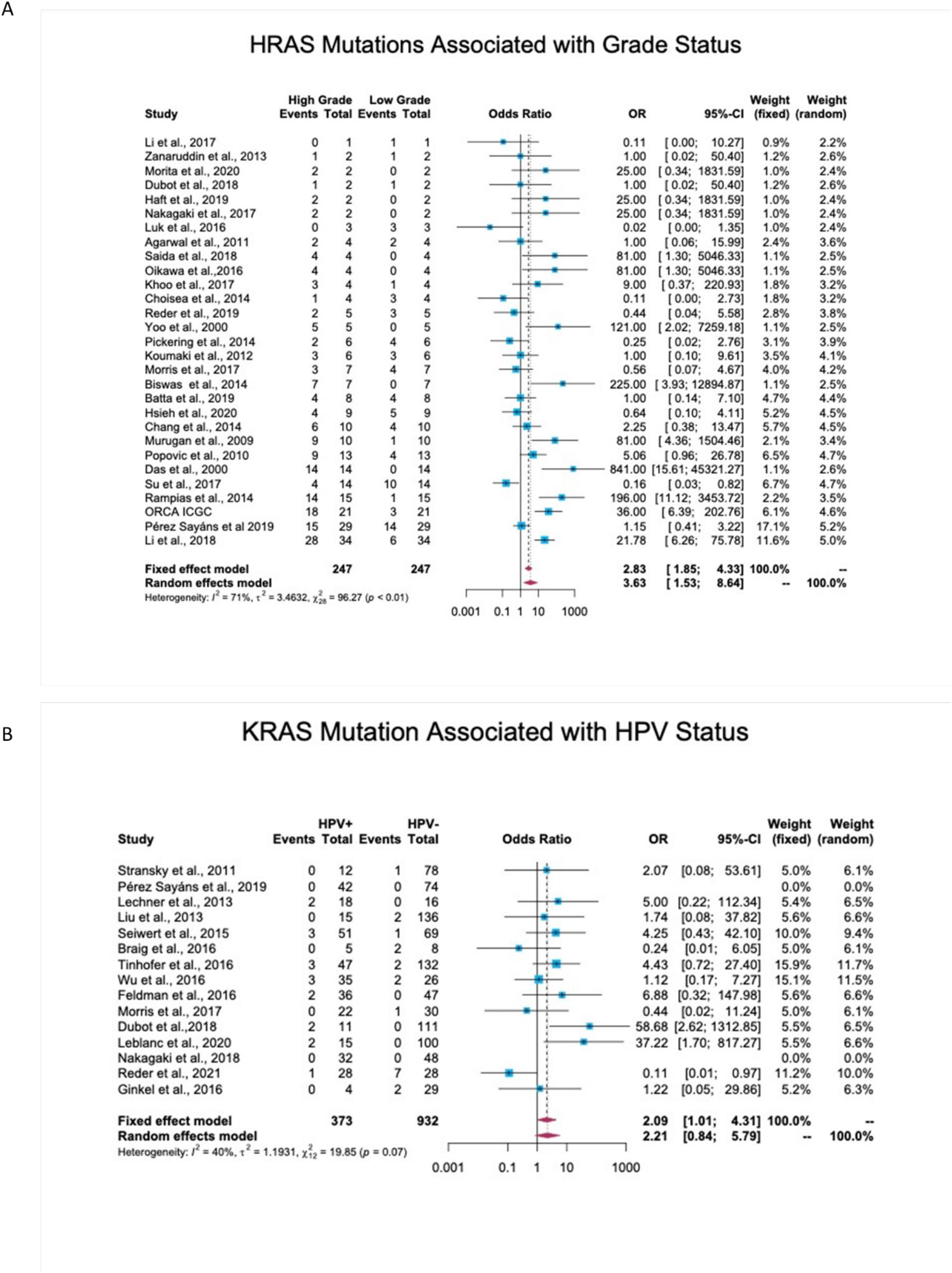
Association between RAS mutations and patient clinical features. **(A)** Association between HRAS mutations and tumor grade. In total, 44 studies reported details regarding tumor stage or grade and mutation status. Stage/grade 1 and 2 tumors were categorized as low-grade, while stage/grade 3 and 4 tumors were categorized as high-grade. An OR analysis exhibited a significant association between HRAS mutation status and advanced stage. (OR = 3.63; 95% CI, 1.53-8.64) **(B)** Association between KRAS mutations and human papillomavirus (HPV) infection status. In total, 17 studies reported the detection of RAS mutations in both HPV-positive and HPV-negative patients. An OR analysis revealed a significant association between KRAS mutation status and HPV infection (OR = 2.09; 95% CI, 1.01-4.31). CI: Confidence interval. I^2^: Inconsistency index. OR: Odds ratio.

### Association between RAS Mutations and HPV Status

Of the 38 cohort studies that reported the HPV status of patients, only 25 provided specific patient data, and of these, 17 included both HPV-negative and HPV-positive patients, thus allowing an OR analysis. This analysis revealed a significant association between HPV-positive status and KRAS mutations, with an OR of 2.09 (95% CI, 1.01-4.31) (Figure 5B), but no significant correlation between HPV-positive status and HRAS or NRAS mutations (eFigure 6 in the Supplement).

## Discussion

After years of extensive research, new strategies that target the RAS-MAPK pathway are now opening new therapeutic options for affected patients.^6^ The meta-analysis presented here compiles findings from the past two decades and provides updated insight into the global prevalence of mutations in RAS family genes, underscoring their potential as therapeutic targets in HNC patients.

The prevalence of mutations was highest for the HRAS gene, followed by KRAS and NRAS. This finding aligns with previous reports of the higher frequency of HRAS mutations in HNC as compared to its frequency in other cancer types in which KRAS mutations are most prevalent, followed by NRAS mutations.^4^ The results of our prevalence analysis diverge slightly from the results of The Cancer Genome Atlas project,^14,15^ which has carried out one of the most significant studies on an HNC patient population. These slight differences may be due to the more heterogeneous population of patients from diverse geographical regions, disease stages, and detection methods included in our analysis.

Our analyses revealed differences in the prevalences of RAS mutations according to the anatomical site, which may account for some of the heterogeneity between cohorts in the overall prevalence analysis. HRAS mutations were more prevalent in oral cavity and salivary gland tumors. In contrast, KRAS mutations were more frequent in sinonasal tumors, and NRAS mutations were found chiefly in tumors of the nasopharynx. These findings emphasize the importance of taking the anatomical site of the tumor into consideration so as to achieve a more accurate assessment of RAS mutation frequencies. The variation in frequencies between tissue types may be due to differences in baseline expression and activity of RAS in different anatomical sites, which may, in turn, affect cellular reprogramming and tumor formation.^4^ Another explanation might be differences in the quality and quantity of exposure to risk factors.^16^

Our data reveal a significantly higher prevalence of HRAS mutations in South Asia, corroborating previous studies on oral cancer in India.^17–20^ Those studies identified region-specific risk factors, such as smoking bidis (cigarettes wrapped in a tendu or temburni leaf),^21,22^ chewing betel nuts,^23^ and oral hygiene,^24^ that contribute, separately or synergistically, to the development of tumors, specifically within the oral cavity.^25–28^ Indeed, in our database, 86% of the patients from South Asia suffered from oral cancer, as opposed to primary tumors in other sites. As noted above, the HRAS mutation frequency is higher in oral cancer worldwide. Thus, further studies are needed to determine whether exposure to such risk factors directly causes mutations in HRAS or whether these factors increase the odds of tumors developing in the oral cavity, in which the prevalence of HRAS mutations is high.

We found that the most frequent amino acid substitution in codon 12 of KRAS was G12D (51%), followed by G12V (16.3%) and G12C (12.9%). This finding provides an indication of the size of the population that could benefit from treatment with mutant-specific inhibitors, i.e., G12C and G12D KRAS inhibitors, that are in various stages of development.

A considerable percentage of HRAS mutations were present in codon 61, particularly in salivary gland tumors. To date, only limited studies have been performed to elucidate the etiology of these specific alterations, but recent analyses for patients with salivary gland cancer have indicated the diagnostic significance of these mutations.^29^ These findings may help evaluate the size of the subpopulations that could benefit from a particular treatment.

Data regarding the association between RAS gene mutations and prognosis in HNC are contradictory. Some studies link RAS mutations with stage and disease recurrence,^30–33^ while others predict better prognosis and overall survival.^34–36^ Our meta-analysis found that mutations in HRAS are significantly associated with high stage/grade scores, emphasizing the importance of considering RAS mutational status when assessing patient prognosis. KRAS and NRAS mutations also exhibited a trend towards being associated with high stage/grade scores. The lower number of cases with KRAS or particularly of NRAS mutations that were available for OR analysis may account for the observed lack of statistical significance.

An association between RAS mutations and HPV status in HNC has been suggested.^31,37^ In accordance with these studies, our data reveal a significant association between HPV-positive status and KRAS mutations. Studies on HPV-related cancers, mainly cervical cancer, demonstrate a similar association.^38–40^ Notably, KRAS mutations, HRAS mutations, and HPV infection were mutually exclusive in benign neoplasms of the head and neck.^41^ These findings suggest that RAS mutations in the context of HPV infection contribute to carcinogenesis.

Several inhibitors of the RAS-MAPK pathway are currently under evaluation as therapeutics for various cancers (reviewed in ^6^). Therefore, knowledge regarding the prevalence of RAS family mutations and associated characteristics in HNC may enable researchers to better assess the need for and the potential of trials with molecularly relevant targeted therapeutics.

### Limitations

Certain methodological limitations of this review should be considered. First, even after selecting only those studies with a low ‘risk of bias score,’ the heterogeneity between studies remained high. We believe that this is due to the heterogeneous nature of HNC, which includes a wide range of anatomical sites and etiologies. We attempted to address this issue by conducting additional sub-group analyses, which consistently revealed significant differences between groups. A second limitation of this analysis derives from the differences in the sequencing methods used in the various studies, which may have influenced overall pooled results by interfering with the accuracy and precision of pooled estimates. Third, we did not analyze sufficient data about patient-specific risk factor exposure. Such data could potentially have strengthened the observed associations in this study and provided additional insights.

## Conclusions

This study highlights RAS as a potential therapeutic target in certain subsets of HNC patients. The findings underscore the differences in the prevalence rates of HRAS, KRAS, and NRAS according to tumor anatomical site and geographical region. The analysis also demonstrates that RAS mutations are associated with tumor stage and HPV status.

## Data Availability

All data produced in the present study are available upon reasonable request to the authors

## Acknowledgments

This work was funded by the Israel Science Foundation (ISF, 302/21 and 700/16) (to M.E.), the Israel Cancer Research Foundation (ICRF, 17-1693-RCDA) (to M.E), United States-Israel Binational Science Foundation (BSF, 2017323) (to M.E). Fellowships: Eileen & Louis Dubrovsky Doctoral Cancer Fellowship Endowment Fund, BGU fellow to O.N.; and a PBC post-doctoral fellowship from Israeli Council for Higher Education to S.J.

