## Supplementary material for "Worldwide Prevalence and Clinical Characteristics of RAS Mutations in Head and Neck Cancer: A Systematic Review and Meta-Analysis": Online Supplementary materials

**Online-Only Supplements**

| Page 1 | eMethods | Search strings |
| --- | --- | --- |
| Page 2 | eFigure 1 | Begg’s funnel plots |
| Pages 3-7 | eTable 1 | Literature references list of studies included in the meta-analysis |
| Pages 8-15 | eTable 2 | Detailed characteristics of the studies included in the meta-analysis |
| Pages 16-31 | eTable 3 | Detailed anatomical site data |
| Page 32 | eFigure 2 | Forest plot of RAS mutation frequency according to geographical region |
| Pages 33-35 | eFigure 3 | Forest plot of RAS mutation frequency according to anatomical site |
| Page 36 | eFigure 4 | Amino Acid Substitutions |
| Page 37 | eFigure 5 | Association between RAS Mutations and Disease Stage/Grade |
| Page 38 | eFigure 6 | Association between RAS Mutations and HPV Status |

**eMethods: Search strings**

A systematic literature review was conducted by searching the PubMed, Embase, Web of Science, and Cochrane Central Register of Controlled Trials databases in June 2021 for studies published in the English language since 1 January 2000. The search string included 'RAS' and ‘mutation’ and one of the following terms: ‘Head and neck cancer’, ‘Head and neck squamous cell carcinoma’, ‘Oral cancer’, ‘oral squamous cell carcinoma’, ‘tongue’, ‘lips’, ‘nasopharyngeal’/’nasopharynx’, ‘pharyngeal’/’pharynx’, ‘laryngeal’/’larynx’, ‘oropharyngeal’/’oropharynx’, ‘Salivary gland’, ‘sinonasal’/’nasal’/’sinus’, ‘oropharyngeal’/’oropharynx’, ‘hypopharyngeal’/’hypopharynx’, or ‘tonsil’.

**eFigure 1: Begg’s funnel plots**

Potential publication bias was analyzed by Begg’s funnel plots, displaying the prevalence of mutations (x-axis) versus the standard error in each study (y-axis). Each dot represents a single study. The white rectangle indicates a 95% pseudo-confidence interval, and the solid middle line indicates the overall effect from the meta-analysis.

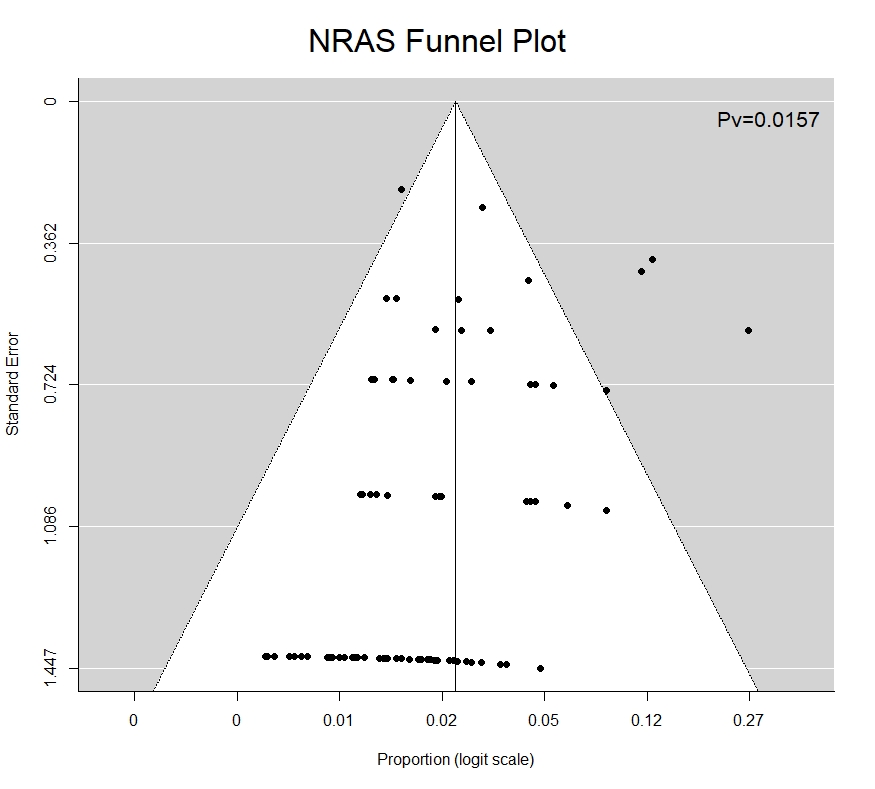

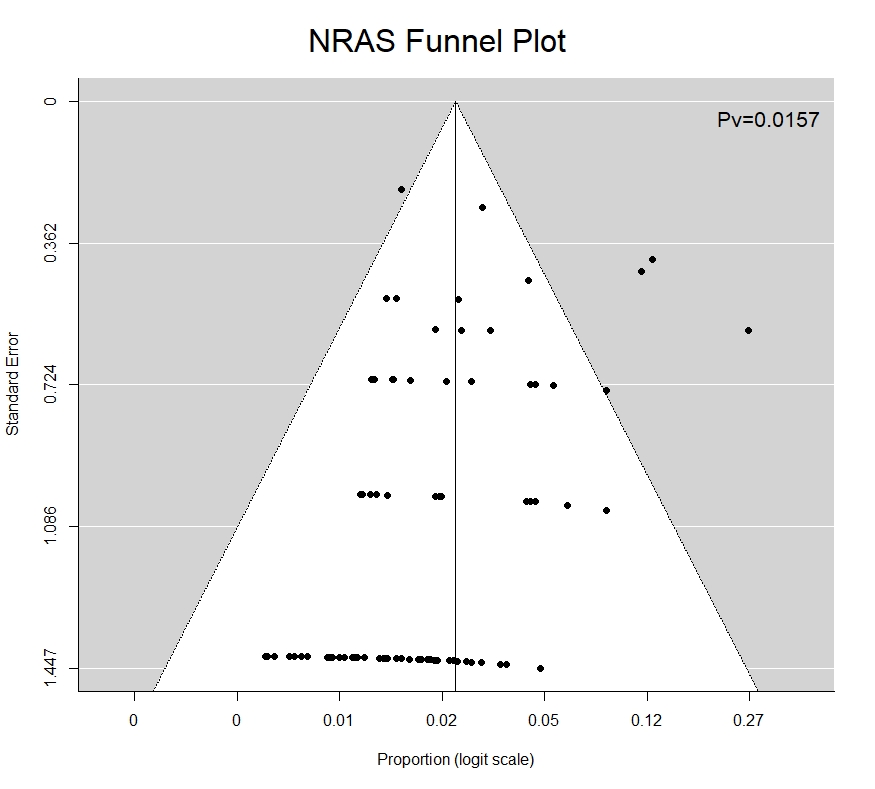

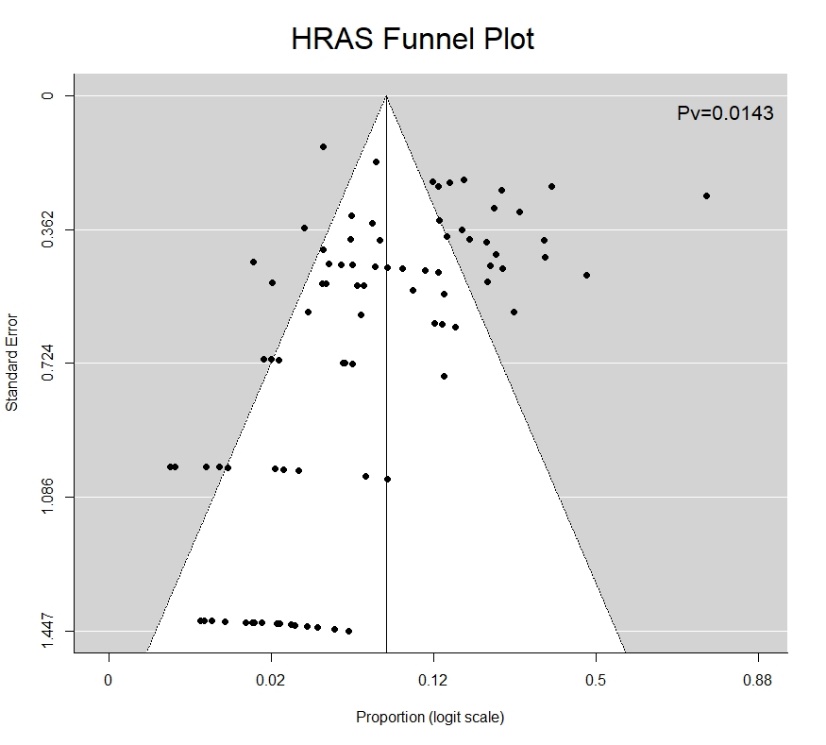

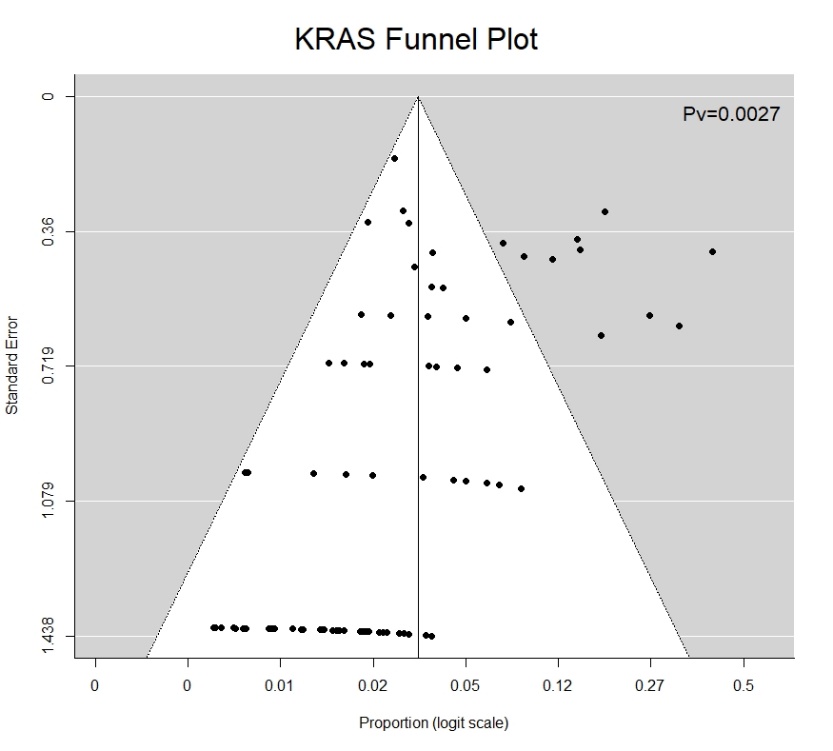

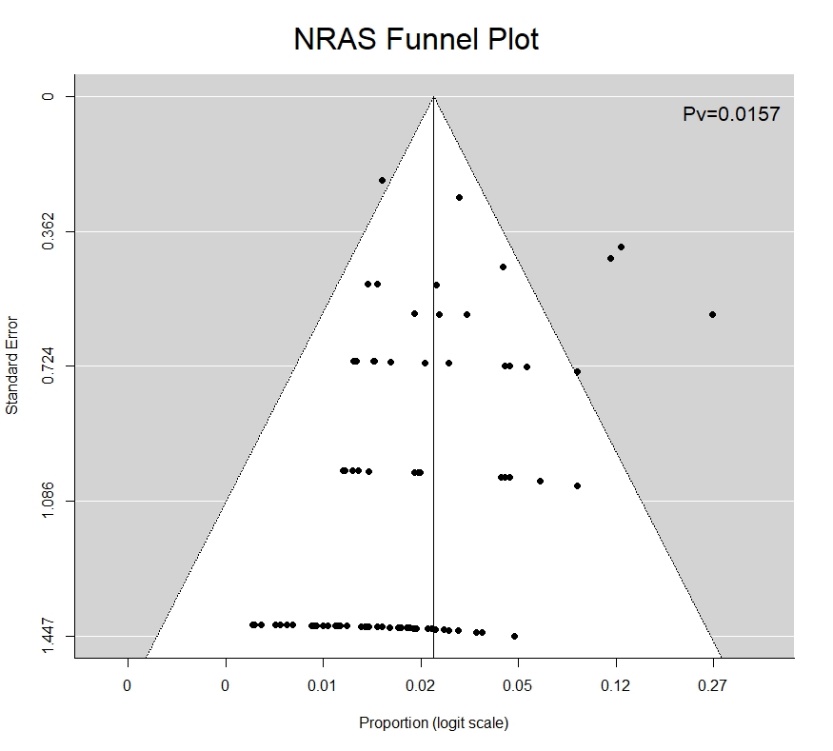

**eTable 1: Literature references list of studies included in the meta-analysis**

[129]	Samstein RM, Lee C-H, Shoushtari AN, Hellmann MD, Shen R, Janjigian YY, et al. Tumor mutational load predicts survival after immunotherapy across multiple cancer types n.d. https://doi.org/10.1038/s41588-018-0312-8.

[135]	Garo Kyurkchiyan S, Kyurkchiyan SG, Popov TM, Kachakova D, Mihova K, Petkova VY, et al. PATHOGENIC SOMATIC ALTERATIONS IN ADVANCED HPV-NEGATIVE CELL SQUAMOUS LARYNGEAL CARCINOMA REVEALED VIA TARGETED NEXT GENERATION SEQUENCING n.d. https://doi.org/10.2298/GENSR2002477K.

**eTable 2: Detailed characteristics of the studies included in the meta-analysis**

|  | **HRAS** | | | **KRAS** | | | **NRAS** | | | **HPV status** | **Sampling technique** |  | | **Risk factors** | **Geographical region** |
| --- | --- | --- | --- | --- | --- | --- | --- | --- | --- | --- | --- | --- | --- | --- | --- |
| **Cohort** | **cases** | **cohort size** | **%** | **cases** | **cohort size** | **%** | **cases** | **cohort size** | **%** |  |  |  |  | |  |
| Das et al., 2000 | 14 | 50 | 28.00 | 4 | 12 | 33.33 | 0 | 50 | 0.00 | NP | PCR+Direct seq |  | T, A, B | | India |
| Yoo et al., 2000 | 5 | 24 | 20.83 | 2 | 24 | 8.33 |  |  |  | NP | PCR+Direct seq |  | NP | | USA |
| Yoo et al., 2000 | 9 | 50 | 18.00 |  |  |  |  |  |  | NP | PCR+Direct seq |  | NP | | USA |
| Lin et al., 2002 | 10 | 28 | 35.71 |  |  |  |  |  |  | NP | PCR+Direct seq |  | NP | | China |
| Weber et al., 2003 |  |  |  | 5 | 89 | 5.62 |  |  |  | NP | PCR+Direct seq |  | NP | | Germany |
| Perrone et al., 2003 | 0 | 18 | 0.00 |  |  |  |  |  |  | NP | PCR+Direct seq |  | W | | Italy |
| Ruiz-Godoy et al., 2006 |  |  |  | 0 | 20 | 0.00 |  |  |  | NP | PCR+Direct seq |  | A, S | | Mexico |
| Sathyan et al., 2006 |  |  |  |  |  |  |  |  |  | NP | PCR-SSCP |  | NP | | India |
| Frattini et al., 2006 |  |  |  | 9 | 18 | 50.00 |  |  |  | NP | PCR+Direct seq |  | W | | Italy |
| Augello et al., 2006 | 11 | 33 | 33.33 | 2 | 33 | 6.06 |  |  |  | NP | SSCP |  | NP | | Italy |
| Russo et al., 2006 | 5 | 81 | 6.17 | 0 | 81 | 0.00 | 0 | 81 | 0.00 | NP | PCR+Direct seq |  | NP | | Italy |
| Sathyan et al., 2007 | 19 | 152 | 12.50 | 0 | 152 | 0.00 | 0 | 152 | 0.00 | NP | PCR+Direct seq |  | T, A | | India |
| Gupta et al., 2007 |  |  |  | 0 | 35 | 0.00 |  |  |  | NP | PCR+SSCP |  | NP | | India |
| Sheikh Ali et al., 2008 |  |  |  | 0 | 91 | 0.00 |  |  |  | NP | PCR+Direct seq |  | NP | | Japan |
| Chou et al., 2008 |  |  |  | 0 | 45 | 0.00 |  |  |  | NP | PCR+Direct seq |  | NP | | Taiwan |
| Bornholdt et al., 2008 |  |  |  | 8 | 174 | 4.60 |  |  |  | NP | PCR+Direct seq |  | W, S, C | | Denmark |
| Murugan et al., 2008 | 3 | 4 | 75.00 |  |  |  |  |  |  | HPV Neg | PCR+Direct seq |  | NP | | India |
| Dahse et al., 2009 |  |  |  | 1 | 65 | 1.54 |  |  |  | NP | PCR+Direct seq |  | NP | | Germany |
| Murugan et al., 2009 | 10 | 56 | 17.86 |  |  |  |  |  |  | NP | PCR+Direct seq |  | T, A | | Vietnam |
| Bruckman et al., 2010 |  |  |  | 1 | 42 | 2.38 |  |  |  | NP | PCR+Direct seq |  | NP | | USA |
| van Damme et al., 2010 |  |  |  | 1 | 22 | 4.55 |  |  |  | NP | PCR+Direct seq |  | NP | | Belgium |
| Chang et al., 2010 | 8 | 58 | 13.79 | 0 | 58 | 0.00 | 0 | 58 | 0.00 | NP | PCR+Direct seq |  | NP | | Taiwan |
| Tetsu et al., 2010 | 8 | 17 | 47.06 | 1 | 17 | 5.88 | 1 | 17 | 5.88 | NP | PCR+Direct seq |  | NP | | USA |
| Murray et al., 2010 |  |  |  | 1 | 92 | 1.09 |  |  |  | NP | PCR+Direct seq |  | T, A | | Greece |
| Popovic et al., 2010 | 13 | 60 | 21.67 |  |  |  |  |  |  | NP | PCR+Direct seq |  | S | | Serbia |
| Agarwal et al., 2011 | 5 | 120 | 4.17 | 0 | 120 | 0.00 | 0 | 120 | 0.00 | Both | WES |  | T, A | | USA |
| Stransky et al., 2011 | 4 | 74 | 5.41 | 1 | 74 | 1.35 | 0 | 74 | 0.00 | Both | WES |  | T, A | | USA |
| Cohen et al., 2011 | 0 | 37 | 0.00 | 0 | 37 | 0.00 | 0 | 37 | 0.00 | NP | MASS SPEC and direct seq |  | NP | | Israel |
| Wang et al., 2011 |  |  |  | 0 | 47 | 0.00 |  |  |  | NP | PCR+Direct seq |  | NP | | Taiwan |
| Friedland et al., 2011 |  |  |  | 0 | 60 | 0.00 |  |  |  | Both | PCR+SSCP |  | S, A | | Australia |
| Szabo et al., 2011 |  |  |  | 2 | 71 | 2.82 |  |  |  | Both | PCR+ RFLP |  | S, A | | Hungary |
| Suda et al., 2012 |  |  |  | 3 | 115 | 2.61 |  |  |  | NP | PCR+Direct seq |  | T | | Japan |
| Smilek et al. 2012 |  |  |  | 4 | 28 | 14.29 |  |  |  | NP | qPCR |  | NP | | Czech republic |
| Koumaki et al., 2012 | 6 | 86 | 6.98 |  |  |  |  |  |  | NP | PCR+Direct seq |  | NP | | Greece |
| Lopez et al., 2012 |  |  |  | 7 | 58 | 12.07 |  |  |  | NP | PCR+Direct seq |  | W, T | | Spain |
| Bissada et al., 2013 |  |  |  | 7 | 195 | 3.59 |  |  |  | NP | PCR+ RFLP |  | NP | | Canada |
| Choisea et al., 2013 | 1 | 58 | 1.72 |  |  |  |  |  |  | HPV Pos | PCR+Direct seq |  | HPV, S | | USA |
| Lechner et al., 2013 | 0 | 34 | 0.00 | 2 | 34 | 5.88 | 0 | 34 | 0.00 | Both | MassArray platform |  | S, A | | UK |
| Pickering et al., 2013 | 5 | 40 | 12.50 | 0 | 40 | 0.00 | 0 | 40 | 0.00 | NP | WES |  | S, A | | USA |
| Ho et al., 2013 | 5 | 59 | 8.47 | 0 | 59 | 0.00 | 1 | 59 | 1.69 | NP | WES |  | NP | | USA |
| Stephens et al., 2013 | 0 | 24 | 0.00 | 0 | 24 | 0.00 | 0 | 24 | 0.00 | NP | WES |  | NP | | UK |
| Liu et al., 2013 | 7 | 151 | 4.64 | 2 | 151 | 1.32 | 0 | 151 | 0.00 | Both | WES |  | NP | | USA |
| Cros et al., 2013 | 7 | 107 | 6.54 | 4 | 107 | 3.74 | 2 | 107 | 1.87 | NP | PCR+Direct seq |  | NP | | France |
| Fuji et al., 2013 |  |  |  | 0 | 183 | 0.00 |  |  |  | NP | PCR+Direct seq |  | S, A | | Japan |
| Projetti et al., 2013 |  |  |  | 2 | 34 | 5.88 |  |  |  | NP | High Resolution Melting Analysis |  | W | | France |
| Szablewski et al., 2013 |  |  |  | 12 | 28 | 42.86 |  |  |  | NP | High Resolution Melting Analysis |  | W, S | | France |
| Carvalho et al., 2013 |  |  |  | 0 | 94 | 0.00 |  |  |  | NP | PCR+Direct seq |  | S, A | | Brazil |
| Zanaruddin et al., 2013 | 3 | 107 | 2.80 | 0 | 107 | 0.00 | 0 | 107 | 0.00 | Both | MassArray platform |  | S, A, B | | Malaysia |
| Wetterskog et al., 2013 | 1 | 48 | 2.08 | 0 | 48 | 0.00 | 0 | 48 | 0.00 | NP | PCR+Direct seq |  | NP | | USA |
| Fury et al., 2013 |  |  |  | 1 | 18 | 5.56 | 0 | 18 | 0.00 | Both | MASS SPEC and direct seq |  | S | | USA |
| Chang et al., 2014 | 10 | 79 | 12.66 | 0 | 79 | 0.00 | 0 | 79 | 0.00 | NP | Primer extension analysis |  | S, A, B | | Taiwan |
| Zhang et al., 2014 | 1 | 53 | 1.89 | 6 | 53 | 11.32 | 6 | 53 | 11.32 | Both | Targeted NGS |  | S, A | | USA |
| Lin et al., 2014 | 0 | 56 | 0.00 | 1 | 56 | 1.79 | 1 | 56 | 1.79 | NP | WES, SNP array |  | EBV | | Singapore |
| Ross et al., 2014 | 0 | 15 | 0.00 | 0 | 15 | 0.00 | 0 | 15 | 0.00 | NP | Targeted NGS |  | NP | | USA |
| Prigge et al., 2014 |  |  |  | 0 | 25 | 0.00 |  |  |  | HPV Positive | PCR+Direct seq |  | NP | | Germany |
| Tan et al., 2014 |  |  |  | 0 | 66 | 0.00 |  |  |  | NP | MassArray platform |  | S | | Singapore |
| Chung et al., 2014 |  |  |  |  |  |  |  |  |  | Both | Micro Array |  | NP | | USA |
| Boeckx et al., 2014 |  |  |  | 1 | 43 | 2.33 |  |  |  | Both | High Resolution Melting Analysis and KRASstrip Assay |  | S | | Belgium |
| Choisea et al., 2014 | 4 | 14 | 28.57 |  |  |  |  |  |  | NP | PCR+Direct seq |  | NP | | USA |
| Lin et al., 2014 |  |  |  | 1 | 66 | 1.52 | 0 | 66 | 0.00 | NP | Targeted NGS |  | EBV | | Singapore |
| Zhang et al., 2014 | 1 | 123 | 0.81 | 0 | 123 | 0.00 | 5 | 123 | 4.07 | NP | MassArray platform |  | NP | | china |
| Franchi et al., 2014 |  |  |  | 1 | 27 | 3.70 |  |  |  | NP | PCR+Direct seq |  | W | | Italy |
| Biswas et al., 2014 | 7 | 84 | 8.33 |  |  |  |  |  |  | Both | WES |  | S, A, B | | India |
| Cortelazzi et al., 2014 | 0 | 54 | 0.00 |  |  |  |  |  |  | Both | PCR+Direct seq |  | NP | | Italy |
| Pickering et al., 2014 | 8 | 39 | 20.51 | 0 | 39 | 0.00 | 2 | 39 | 5.13 | NP | WES |  | NP | | USA |
| Rampias et al., 2014 | 17 | 180 | 9.44 |  |  |  |  |  |  | NP | PCR+Direct seq |  | S, A | | Greece |
| Zhang et al., 2015 |  |  |  | 1 | 70 | 1.43 |  |  |  | NP | SnapShot Multiplex assay |  | S, EBV | | china |
| Seiwert et al., 2015 | 4 | 120 | 3.33 | 4 | 120 | 3.33 | 1 | 120 | 0.83 | Both | Targeted NGS |  | S, A | | USA |
| Vettore et al., 2015 | 1 | 60 | 1.67 |  |  |  |  |  |  | NP | Targeted NGS |  | T | | Singapore |
| Chen et al., 2015 | 32 | 345 | 9.28 | 8 | 345 | 2.32 | 4 | 345 | 1.15 | NP | Targeted NGS |  | S, A, B | | Taiwan |
| Fu et al., 2015 |  |  |  | 2 | 18 | 11.11 | 0 | 18 | 0.00 | NP | MassArray platform |  | NP | | Taiwan |
| Grunewald et al., 2015 | 20 | 84 | 23.81 | 0 | 84 | 0.00 | 2 | 84 | 2.38 | NP | Targeted NGS |  | NP | | Germany |
| Kato et al., 2015 | 13 | 117 | 11.11 | 4 | 117 | 3.42 |  |  |  | NP | Targeted NGS |  | NP | | USA |
| Choisea et al., 2015 | 10 | 29 | 34.48 | 0 | 29 | 0.00 | 0 | 29 | 0.00 | NP | Targeted NGS |  | NP | | USA |
| Er et al., 2015 |  |  |  | 0 | 50 | 0.00 | 0 | 50 |  | NP | Targeted NGS |  | S, A, B | | Taiwan |
| Fonseca et al., 2015 | 0 | 17 | 0.00 | 3 | 17 | 17.65 | 1 | 17 | 5.88 | NP | Targeted NGS |  | NP | | USA |
| Braig et al., 2016 | 5 | 46 | 10.87 | 2 | 46 | 4.35 | 2 | 46 | 4.35 | Both | Targeted NGS |  | NP | | Germany |
| Rettig et al., 2016 | 0 | 25 | 0.00 | 0 | 25 | 0.00 | 1 | 25 | 4.00 | NP | WGS |  | NP | | USA |
| Mitani et al., 2016 | 0 | 65 | 0.00 | 0 | 65 | 0.00 | 0 | 65 | 0.00 | NP | WGS |  | NP | | USA |
| Drier et al., 2016 | 0 | 10 | 0.00 | 0 | 19 | 0.00 | 0 | 10 | 0.00 | NP | WGS |  | NP | | USA |
| Shalmon et al., 2016 |  |  |  | 0 | 21 | 0.00 |  |  |  | NP | PCR+Direct seq |  | NP | | Israel |
| Wang et al., 2016 | 21 | 149 | 14.09 | 0 | 149 | 0.00 | 0 | 149 | 0.00 | NP | Targeted NGS |  | NP | | USA |
| Tinhofer et al., 2016 | 1 | 179 | 0.56 | 5 | 179 | 2.79 | 2 | 179 | 1.12 | HPV Pos | Targeted NGS |  | S | | Germany |
| Schneider et al., 2016 |  |  |  | 0 | 43 | 0.00 |  |  |  | NP | PCR+Direct seq |  | NP | | Germany |
| Choisea et al., 2016 | 13 | 38 | 34.21 |  |  |  |  |  |  | NP | Targeted NGS |  | NP | | USA |
| Dalin et al., 2016 | 7 | 31 | 22.58 | 0 | 31 | 0.00 | 0 | 31 | 0.00 | NP | WES |  | NP | | USA |
| Wu et al., 2016 | 8 | 214 | 3.74 | 18 | 214 | 8.41 |  |  |  | NP | MassArray platform |  | NP | | USA |
| Chau et al., 2016 | 10 | 213 | 4.69 | 1 | 213 | 0.47 | 2 | 213 | 0.94 | NP | Targeted NGS |  | NP | | USA |
| Bell et al., 2016 | 0 | 24 | 0.00 | 10 | 24 | 41.67 | 2 | 24 | 8.33 | NP | MassArray platform |  | NP | | USA |
| Luk et al., 2016 | 3 | 23 | 13.04 | 0 | 23 | 0.00 | 1 | 23 | 4.35 | NP | MassArray platform |  | NP | | Australia |
| Al-Hebshi et al., 2016 | 3 | 20 | 15.00 | 0 | 20 | 0.00 | 0 | 20 | 0.00 | HPV Neg | WES |  | S, EBV | | Yemen |
| Hedberg et al., 2016 | 0 | 12 | 0.00 | 1 | 12 | 8.33 | 1 | 12 | 8.33 | NP | WES |  | NP | | USA |
| Feldman et al., 2016 | 8 | 299 | 2.68 | 11 | 448 | 2.46 | 4 | 381 | 1.05 | Both | sanger and Targeted NGS |  | NP | | USA |
| Ock et al., 2016 | 26 | 71 | 36.62 | 13 | 71 | 18.31 | 20 | 71 | 28.17 | NP | Targeted NGS |  | S | | Korea |
| Oikawa et al.,2016 | 4 | 220 | 1.82 | 1 | 220 | 0.45 | 2 | 220 | 0.91 | NP | Targeted NGS |  | NP | | Japan |
| Ginkel et al., 2016 | 10 | 110 | 9.09 | 2 | 110 | 1.82 | 0 | 110 | 0.00 | Both | Targeted NGS |  | S, A | | Netherlands |
| Morris et al., 2017 | 9 | 151 | 5.96 | 2 | 151 | 1.32 | 2 | 151 | 1.32 | Both | Targeted NGS |  | S, A | | USA |
| Zhang et al., 2017 | 2 | 94 | 2.13 | 3 | 94 | 3.19 | 1 | 94 | 1.06 | EBV pos | WES |  | S, EBV | | China |
| Ali et al., 2017 | 1 | 190 | 0.53 | 3 | 190 | 1.58 | 4 | 190 | 2.11 | Not provided | Targeted NGS |  | EBV | | China |
| Upadhyay et al., 2017 | 3 | 25 | 12.00 | 0 | 25 | 0.00 | 0 | 25 | 0.00 | NP | WES |  | S, B | | India |
| Kang et al., 2017 | 1 | 18 | 5.56 | 0 | 18 | 0.00 | 0 | 18 | 0.00 | NP | WES |  | T | | USA |
| Dalin et al., 2017 | 1 | 40 | 2.50 | 0 | 40 | 0.00 | 0 | 40 | 0.00 | NP | WES |  | NP | | USA |
| Li et al., 2017 | 2 | 105 | 1.90 | 1 | 105 | 0.95 | 4 | 105 | 3.81 | NP | WES |  | NP | | USA/Hong Kong |
| Dogan et al., 2017 | 0 | 30 | 0.00 | 3 | 40 | 7.50 | 0 | 30 | 0.00 | NP | Targeted NGS |  | NP | | USA |
| Zehir et al., 2017 | 11 | 105 | 10.48 | 3 | 105 | 2.86 | 4 | 105 | 3.81 | NP | Targeted NGS |  | NP | | USA |
| Bersani et al., 2017 | 5 | 344 | 1.45 | 9 | 344 | 2.62 | 4 | 344 | 1.16 | Both | Targeted NGS |  | S | | Sweden |
| Nakagaki et al., 2017 | 2 | 47 | 4.26 | 0 | 47 | 0.00 | 0 | 47 | 0.00 | NP | Targeted NGS |  | NP | | Japan |
| Su et al., 2017 | 14 | 120 | 11.67 |  |  |  |  |  |  | NP | WES |  | S, A, B | | Taiwan |
| Robinson et al., 2017 | 3 | 15 | 20.00 | 1 | 15 | 6.67 | 0 | 15 | 0.00 | NP | WES |  | NP | | USA |
| Khoo et al., 2017 | 4 | 15 | 26.67 | 4 | 14 | 28.57 | 4 | 15 | 26.67 | NP | Targeted NGS |  | NP | | Australia |
| Saida et al., 2018 | 4 | 70 | 5.71 | 6 | 70 | 8.57 | 0 | 70 | 0.00 | NP | Micro Array |  | NP | | Japan |
| Hallani et al., 2018 | 8 | 23 | 34.78 | 0 | 23 | 0.00 | 0 | 23 | 0.00 | NP | Targeted NGS |  | NP | | USA |
| Shimura et al., 2018 | 23 | 140 | 16.43 | 0 | 140 | 0.00 | 0 | 140 | 0.00 | NP | PCR+Direct seq |  | NP | | Multi centered |
| Perdomo et al., 2018 | 20 | 180 | 11.11 |  |  |  |  |  |  | Both | Targeted NGS |  | S, A, HPV | | Europe and South America |
| Vossen etal., 2018 | 2 | 111 | 1.80 | 0 | 111 | 0.00 | 1 | 111 | 0.90 | HPV neg | Targeted NGS |  | S, A | | Netherlands |
| Dubot et al.,2018 | 2 | 122 | 1.64 | 2 | 122 | 1.64 | 1 | 122 | 0.82 | Both | PCR+Direct seq |  | S, A, HPV | | France |
| Mirghani et al., 2018 | 0 | 62 | 0.00 | 2 | 62 | 3.23 | 0 | 62 | 0.00 | HPV Pos | Targeted NGS |  | S, T, HPV | | France |
| Nakagaki et al., 2018 | 4 | 80 | 5.00 | 0 | 80 | 0.00 | 0 | 80 | 0.00 | Both | Targeted NGS |  | NP | | Japan |
| Li et al., 2018 | 34 | 168 | 20.24 |  |  |  |  |  |  | NP | Amplification-refractory mutation system |  | NP | | China |
| Batta et al., 2019 | 8 | 46 | 17.39 | 0 | 46 | 0.00 | 0 | 46 | 0.00 | NP | Targeted NGS |  | T | | India |
| Akagi et al., 2019 |  |  |  | 1 | 85 | 1.18 |  |  |  | NP | PCR+Direct seq |  | NP | | Japan |
| Chung et al., 2019 | 0 | 33 | 0.00 | 1 | 33 | 3.03 | 0 | 33 | 0.00 | NP | Targeted NGS |  | S, EBV | | Taiwan |
| Reder et al., 2019 | 5 | 12 | 41.67 | 1 | 12 | 8.33 | 1 | 12 | 8.33 | NP | Targeted NGS |  | S, A | | Germany |
| Stanek et al., 2019 |  |  |  | 7 | 55 | 12.73 | 2 | 54 | 3.70 | Both | PCR- mutation detection kit |  | NP | | Czech republic |
| Nakaguro et al., 2019 | 7 | 21 | 33.33 |  |  |  |  |  |  | NP | PCR+Direct seq |  | NP | | Japan |
| Wang et al., 2019 |  |  |  | 30 | 80 | 37.50 |  |  |  | NP | PCR+Direct seq |  | S | | China |
| Haft et al., 2019 | 2 | 46 | 4.35 | 0 | 46 | 0.00 | 0 | 46 | 0.00 | HPV Pos | WES |  | S, A, HPV | | USA |
| Smith et al., 2019 | 0 | 21 | 0.00 | 1 | 21 | 4.76 | 0 | 21 | 0.00 | NP | Targeted NGS |  | S | | USA |
| Westbrook et al., 2019 |  |  |  | 1 | 23 | 4.35 | 0 | 23 | 0.00 | NP | Targeted NGS |  | S | | USA |
| Samstein et al., 2019 | 5 | 139 | 3.60 | 3 | 139 | 2.16 | 3 | 139 | 2.16 | NP | Targeted NGS |  | NP | | USA |
| Priestley et al., 2019 | 2 | 42 | 4.76 | 0 | 42 | 0.00 | 0 | 42 | 0.00 | NP | WGS |  | NP | | Netherlands |
| Pérez Sayáns et al 2019 | 33 | 528 | 6.25 | 9 | 528 | 1.70 | 14 | 528 | 2.65 | Both | Targeted NGS, WGS, WES |  | S | | Multi centered |
| Kobyashi et al., 2019 | 7 | 284 | 2.46 |  |  |  |  |  |  | Both | PCR+Direct seq |  | S, A | | Japan |
| Dogan et al., 2019 | 6 | 25 | 24.00 | 0 | 25 | 0.00 | 0 | 25 | 0.00 | NP | Targeted NGS |  | S | | USA |
| Gauthaman et al., 2020 |  |  |  | 34 | 56 | 60.71 |  |  |  | NP | PCR-RFLP |  | NP | | India |
| Kyurkchiyan et al., 2020 | 3 | 57 | 5.26 | 2 | 57 | 3.51 | 1 | 57 | 1.75 | HPV Neg | Targeted NGS |  | NP | | Bulgaria |
| Morita et al., 2020 | 2 | 101 | 1.98 | 7 | 101 | 6.93 | 0 | 101 | 0.00 | NP | SnapShot Multiplex assay |  | NP | | Japan |
| Morfouace et al., 2020 | 0 | 14 | 0.00 | 0 | 14 | 0.00 |  |  |  | NP | Targeted NGS |  | NP | | France |
| Kawamura et al., 2020 | 0 | 20 | 0.00 | 0 | 20 | 0.00 | 0 | 20 | 0.00 | NP | PCR-SSCP |  | NP | | Japan |
| Jayaprakash et al., 2020 | 6 | 28 | 21.43 | 0 | 28 | 0.00 | 0 | 28 | 0.00 | NP | Targeted NGS |  | NP | | India |
| Leblanc et al., 2020 | 4 | 115 | 3.48 | 2 | 115 | 1.74 | 0 | 115 | 0.00 | Both | High Resolution Melting Analysis and sanger |  | NP | | France |
| Mueller et al., 2020 | 14 | 63 | 22.22 | 3 | 63 | 4.76 | 0 | 63 | 0.00 | NP | Targeted NGS |  | S | | Australia |
| Kim et al., 2020 | 4 | 42 | 9.52 | 0 | 42 | 0.00 | 0 | 42 | 0.00 | NP | Targeted NGS |  | S | | Korea |
| Hsieh et al., 2020 | 9 | 33 | 27.27 |  |  |  | 8 | 33 | 24.24 | NP | PCR+Direct seq |  | NP | | Taiwan |
| ORCA ICGC | 21 | 178 | 11.80 | 6 | 178 | 3.37 | 2 | 178 | 1.12 | NP | targeted NGS, WGS |  | S, A, B | | India |
| Masato et al., 2021 | 66 | 83 | 79.52 | 0 | 83 | 0.00 | 0 | 83 | 0.00 | NP | PCR+Direct seq |  | NP | | Japan |
| Sanchez-Fernandez et al., 2021 | 0 | 48 | 0.00 | 7 | 48 | 14.58 | 2 | 48 | 4.17 | NP | Targeted NGS |  | S, A, W | | Spain |
| AACR GENIE V9.1 | 55 | 1636 | 3.36 | 37 | 1636 | 2.26 | 20 | 1636 | 1.22 | NP | WES |  | S | | Multi centered |
| Reder et al., 2021 | 9 | 56 | 16.07 | 8 | 56 | 14.29 | 7 | 56 | 12.50 | Both | Targeted NGS |  | S, A | | Germany |
| Patel et al., 2021 | 4 | 30 | 13.33 | 0 | 30 | 0.00 | 0 | 30 | 0.00 | NP | WES |  | T | | India |

NP-not provided, HPV pos- human papillomavirus positive, HPV neg- human papillomavirus negative, S-smoking T-Tabacco, A-Alcohol, B-betel chewing, W wood/leather dust, C- other chemical exposure, HPV-. human papillomavirus, EBV- Epstein–Barr virus

**eTable 3: Detailed anatomical site data**

|  | **HRAS** | | | | | | | | | | | | | | | |
| --- | --- | --- | --- | --- | --- | --- | --- | --- | --- | --- | --- | --- | --- | --- | --- | --- |
|  | **Oral cavity** | | **Salivary gland** | | **sinonasal** | | **nasopharynx** | | **oropharynx** | | **hypopharynx** | | **larynx** | | **other** | |
| **cohort** | mutated | total | mutated | total | mutated | total | mutated | total | mutated | total | mutated | total | mutated | total | mutated | total |
| Das et al., 2000 | 14 | 50 |  |  |  |  |  |  |  |  |  |  |  |  |  |  |
| Yoo et al., 2000 |  |  | 5 | 24 |  |  |  |  |  |  |  |  |  |  |  |  |
| Yoo et al., 2000 |  |  | 9 | 50 |  |  |  |  |  |  |  |  |  |  |  |  |
| Weber et al., 2003 |  |  |  |  |  |  |  |  |  |  |  |  |  |  |  |  |
| Perrone et al., 2003 |  |  |  |  | 0 | 18 |  |  |  |  |  |  |  |  |  |  |
| Ruiz-Godoy et al., 2006 |  |  |  |  |  |  |  |  |  |  |  |  |  |  |  |  |
| Sathyan et al., 2006 |  |  |  |  |  |  |  |  |  |  |  |  |  |  |  |  |
| Frattini et al., 2006 |  |  |  |  |  |  |  |  |  |  |  |  |  |  |  |  |
| Augello et al., 2006 |  |  | 11 | 33 |  |  |  |  |  |  |  |  |  |  |  |  |
| Sathyan et al., 2007 | 19 | 152 |  |  |  |  |  |  |  |  |  |  |  |  |  |  |
| Gupta et al., 2007 | 0 | 20 |  |  |  |  | 0 | 1 |  |  |  |  | 0 | 14 |  |  |
| Sheikh Ali et al., 2008 |  |  |  |  |  |  |  |  |  |  |  |  |  |  |  |  |
| Chou et al., 2008 |  |  |  |  |  |  |  |  |  |  |  |  |  |  |  |  |
| Bornholdt et al., 2008 |  |  |  |  |  |  |  |  |  |  |  |  |  |  |  |  |
| Dahse et al., 2009 |  |  |  |  |  |  |  |  |  |  |  |  |  |  |  |  |
| Murugan et al., 2009 | 10 | 56 |  |  |  |  |  |  |  |  |  |  |  |  |  |  |
| Bruckman et al., 2010 |  |  |  |  |  |  |  |  |  |  |  |  |  |  |  |  |
| van Damme et al., 2010 |  |  |  |  |  |  |  |  |  |  |  |  |  |  |  |  |
| Chang et al., 2010 | 8 | 58 |  |  |  |  |  |  |  |  |  |  |  |  |  |  |
| Tetsu et al., 2010 | 7 | 8 | 0 | 3 | 0 | 1 |  |  |  |  |  |  |  |  | 3 | 5 |
| Murray et al., 2010 |  |  |  |  |  |  |  |  |  |  |  |  |  |  |  |  |
| Agarwal et al., 2011 | 4 | 75 |  |  |  |  |  |  | 1 | 21 | 0 | 9 | 0 | 13 |  |  |
| Stransky et al., 2011 | 3 | 51 |  |  | 0 | 2 |  |  | 0 | 15 | 0 | 9 | 1 | 15 |  |  |
| Cohen et al., 2011 | 0 | 37 |  |  |  |  |  |  |  |  |  |  |  |  |  |  |
| Trivedi et al., 2011 |  |  |  |  |  |  |  |  |  |  |  |  |  |  |  |  |
| Wang et al., 2011 |  |  |  |  |  |  |  |  |  |  |  |  |  |  |  |  |
| Friedland et al., 2011 |  |  |  |  |  |  |  |  |  |  |  |  |  |  |  |  |
| Szabo et al., 2011 |  |  |  |  |  |  |  |  |  |  |  |  |  |  |  |  |
| Szanyi et al., 2011 |  |  |  |  |  |  |  |  |  |  |  |  |  |  |  |  |
| Suda et al., 2012 |  |  |  |  |  |  |  |  |  |  |  |  |  |  |  |  |
| Koumaki et al., 2012 | 3 | 86 |  |  |  |  |  |  |  |  |  |  |  |  |  |  |
| Lopez et al., 2012 and Garcia-Inclan et al., 2012 |  |  |  |  |  |  |  |  |  |  |  |  |  |  |  |  |
| Smilek et al., 2012 |  |  |  |  |  |  |  |  |  |  |  |  |  |  |  |  |
| Bissada et al., 2013 |  |  |  |  |  |  |  |  |  |  |  |  |  |  |  |  |
| Choisea et al., 2013 |  |  |  |  |  |  |  |  | 1 | 58 |  |  |  |  |  |  |
| Lechner et al., 2013 |  |  |  |  |  |  |  |  | 0 | 34 |  |  |  |  |  |  |
| Pickering et al., 2013 | 5 | 40 |  |  |  |  |  |  |  |  |  |  |  |  |  |  |
| Ho et al., 2013 |  |  | 5 | 59 |  |  |  |  |  |  |  |  |  |  |  |  |
| Stephens et al., 2013 |  |  | 0 | 24 |  |  |  |  |  |  |  |  |  |  |  |  |
| Liu et al., 2013 |  |  |  |  |  |  |  |  |  |  |  |  |  |  |  |  |
| Cros et al., 2013 |  |  | 7 | 107 |  |  |  |  |  |  |  |  |  |  |  |  |
| Fuji et al., 2013 |  |  |  |  |  |  |  |  |  |  |  |  |  |  |  |  |
| Projetti et al., 2013 |  |  |  |  |  |  |  |  |  |  |  |  |  |  |  |  |
| Szablewski et al., 2013 |  |  |  |  |  |  |  |  |  |  |  |  |  |  |  |  |
| Carvalho et al., 2013 |  |  |  |  |  |  |  |  |  |  |  |  |  |  |  |  |
| Zanaruddin et al., 2013 | 3 | 107 |  |  |  |  |  |  |  |  |  |  |  |  |  |  |
| Wetterskog et al., 2013 |  |  |  |  |  |  |  |  |  |  |  |  |  |  |  |  |
| Fury et al., 2013 |  |  |  |  |  |  |  |  |  |  |  |  |  |  |  |  |
| Chang et al., 2014 | 10 | 79 |  |  |  |  |  |  |  |  |  |  |  |  |  |  |
| Zhang et al., 2014 |  |  |  |  |  |  |  |  |  |  |  |  |  |  |  |  |
| Lin et al., 2014 |  |  |  |  |  |  | 0 | 56 |  |  |  |  |  |  |  |  |
| Ross et al., 2014 |  |  | 0 | 15 |  |  |  |  |  |  |  |  |  |  |  |  |
| Al Rawi et al., 2014 |  |  |  |  |  |  |  |  |  |  |  |  |  |  |  |  |
| Prigge et al., 2014 |  |  |  |  |  |  |  |  |  |  |  |  |  |  |  |  |
| Tan et al., 2014 |  |  |  |  |  |  |  |  |  |  |  |  |  |  |  |  |
| Chung et al., 2014 |  |  |  |  |  |  |  |  |  |  |  |  |  |  |  |  |
| Boeckx et al., 2014 |  |  |  |  |  |  |  |  |  |  |  |  |  |  |  |  |
| Choisea et al., 2014 |  |  | 4 | 14 |  |  |  |  |  |  |  |  |  |  |  |  |
| Lin et al., 2014 |  |  |  |  |  |  |  |  |  |  |  |  |  |  |  |  |
| Zhang et al., 2014 |  |  |  |  |  |  | 1 | 123 |  |  |  |  |  |  |  |  |
| Franchi et al., 2014 |  |  |  |  |  |  |  |  |  |  |  |  |  |  |  |  |
| Zhang et al., 2015 |  |  |  |  |  |  |  |  |  |  |  |  |  |  |  |  |
| Seiwert et al., 2015 |  |  |  |  |  |  |  |  |  |  |  |  |  |  |  |  |
| Vettore et al., 2015 | 1 | 60 |  |  |  |  |  |  |  |  |  |  |  |  |  |  |
| Chen et al., 2015 | 32 | 345 |  |  |  |  |  |  |  |  |  |  |  |  |  |  |
| Fu et al., 2015 |  |  |  |  |  |  |  |  |  |  |  |  |  |  |  |  |
| Grunewald et al., 2015 |  |  | 20 | 84 |  |  |  |  |  |  |  |  |  |  |  |  |
| Kato et al., 2015 |  |  | 13 | 117 |  |  |  |  |  |  |  |  |  |  |  |  |
| Choisea et al., 2015 |  |  | 10 | 29 |  |  |  |  |  |  |  |  |  |  |  |  |
| Braig et al., 2016 | 3 | 12 |  |  | 0 | 2 |  |  | 1 | 19 | 1 | 9 | 0 | 4 |  |  |
| Rettig et al., 2016 |  |  | 0 | 25 |  |  |  |  |  |  |  |  |  |  |  |  |
| Mitani et al., 2016 |  |  | 0 | 65 |  |  |  |  |  |  |  |  |  |  |  |  |
| Drier et al., 2016 |  |  | 0 | 10 |  |  |  |  |  |  |  |  |  |  |  |  |
| Shalmon et al., 2016 |  |  |  |  |  |  |  |  |  |  |  |  |  |  |  |  |
| Wang et al., 2016 |  |  | 2 | 149 |  |  |  |  |  |  |  |  |  |  |  |  |
| Tinhofer et al., 2016 |  |  |  |  |  |  |  |  |  |  |  |  |  |  |  |  |
| Kucuk et al., 2016 |  |  |  |  |  |  |  |  |  |  |  |  |  |  |  |  |
| Schneider et al., 2016 |  |  |  |  |  |  |  |  |  |  |  |  |  |  |  |  |
| Choisea et al., 2016 |  |  | 13 | 38 |  |  |  |  |  |  |  |  |  |  |  |  |
| Dalin et al., 2016 |  |  | 7 | 31 |  |  |  |  |  |  |  |  |  |  |  |  |
| Udager et al., 2016 |  |  |  |  |  |  |  |  |  |  |  |  |  |  |  |  |
| Wu et al., 2016 | 4 | 56 |  |  |  |  |  |  | 0 | 76 |  |  | 4 | 82 |  |  |
| Chau et al., 2016 | 3 | 60 |  |  | 0 | 5 | 0 | 7 | 3 | 97 | 0 | 6 | 1 | 24 | 3 | 14 |
| Bell et al., 2016 |  |  |  |  |  |  |  |  |  |  |  |  |  |  | 0 | 24 |
| Luk et al., 2016 |  |  | 3 | 23 |  |  |  |  |  |  |  |  |  |  |  |  |
| Al-Hebshi et al., 2016 |  |  |  |  |  |  |  |  |  |  |  |  |  |  |  |  |
| Hedberg et al., 2016 | 0 | 6 |  |  |  |  |  |  |  |  | 0 | 3 | 0 | 4 |  |  |
| Chuerduangphui et al., 2017 |  |  |  |  |  |  |  |  |  |  |  |  |  |  |  |  |
| Morris et al., 2017 | 1 | 26 | 4 | 32 | 1 | 15 | 0 | 9 | 0 | 23 | 0 | 2 | 0 | 8 | 3 | 36 |
| Yue et al., 2017 |  |  |  |  |  |  |  |  |  |  |  |  |  |  |  |  |
| Abdolkarim Moazeni-Roodi et al., 2017 |  |  |  |  |  |  |  |  |  |  |  |  |  |  |  |  |
| Zhang et al., 2017 |  |  |  |  |  |  | 1 | 94 |  |  |  |  |  |  |  |  |
| Ali et al., 2017 |  |  |  |  |  |  |  |  |  |  |  |  |  |  |  |  |
| Upadhyay et al., 2017 | 3 | 24 |  |  |  |  |  |  |  |  |  |  |  |  |  |  |
| kang et al., 2017 |  |  | 1 | 18 |  |  |  |  |  |  |  |  |  |  |  |  |
| Dalin et al., 2017 |  |  | 1 | 40 |  |  |  |  |  |  |  |  |  |  |  |  |
| Li et al., 2017 |  |  |  |  |  |  | 1 | 105 |  |  |  |  |  |  |  |  |
| Dogan et al., 2017 |  |  |  |  | 0 | 30 |  |  |  |  |  |  |  |  |  |  |
| Krishna et al., 2018 |  |  |  |  |  |  |  |  |  |  |  |  |  |  |  |  |
| Lin et al., 2018 |  |  |  |  |  |  |  |  |  |  |  |  |  |  |  |  |
| Saida et al., 2018 |  |  | 4 | 70 |  |  |  |  |  |  |  |  |  |  |  |  |
| Hallani et al., 2018 |  |  | 8 | 23 |  |  |  |  |  |  |  |  |  |  |  |  |
| Shimura et al., 2018 |  |  | 23 | 140 |  |  |  |  |  |  |  |  |  |  |  |  |
| Perdomo et al., 2018 |  |  |  |  |  |  |  |  |  |  |  |  |  |  |  |  |
| Vossen etal., 2018 |  |  |  |  |  |  |  |  |  |  |  |  |  |  |  |  |
| Batta et al., 2019 | 8 | 39 |  |  |  |  |  |  |  |  |  |  |  |  | 0 | 7 |
| Akagi et al., 2019 |  |  |  |  |  |  |  |  |  |  |  |  |  |  |  |  |
| Chung et al., 2019 |  |  |  |  |  |  |  |  |  |  |  |  |  |  |  |  |
| Reder et al., 2019 |  |  |  |  |  |  |  |  | 5 | 12 |  |  |  |  |  |  |
| Stanek et al., 2019 |  |  |  |  |  |  |  |  |  |  |  |  |  |  |  |  |
| Urano et al., 2019 |  |  |  |  |  |  |  |  |  |  |  |  |  |  |  |  |
| Nakaguro et al., 2019 |  |  | 7 | 21 |  |  |  |  |  |  |  |  |  |  |  |  |
| Wang et al., 2019 |  |  |  |  |  |  |  |  |  |  |  |  |  |  |  |  |
| Reder et al., 2019 |  |  |  |  |  |  |  |  | 0 | 12 |  |  |  |  |  |  |
| Gauthaman et al., 2020 |  |  |  |  |  |  |  |  |  |  |  |  |  |  |  |  |
| Kyurkchiyan et al., 2020 |  |  |  |  |  |  |  |  |  |  |  |  |  |  |  |  |
| ORCA ICGC |  |  |  |  |  |  |  |  |  |  |  |  | 3 | 57 |  |  |
| Sasaki et al., 2020 | 6 | 12 |  |  |  |  |  |  |  |  | 7 | 20 | 0 | 7 | 4 | 12 |
| Morita et al., 2020 |  |  | 2 | 101 |  |  |  |  |  |  |  |  |  |  |  |  |
| Morfouace et al., 2020 |  |  |  |  | 0 | 3 | 0 | 10 |  |  |  |  |  |  | 0 | 1 |
| Kawamura et al., 2020 | 0 | 20 |  |  |  |  |  |  |  |  |  |  |  |  |  |  |
| Pérez Sayáns et al 2019 | 28 | 303 |  |  |  |  |  |  | 1 | 82 | 0 | 10 | 1 | 117 |  |  |
| Masato et al., 2021 |  |  |  |  |  |  |  |  |  |  |  |  |  |  |  |  |
| Sanchez-Fernandez et al., 2021 |  |  |  |  | 0 | 48 |  |  |  |  |  |  |  |  |  |  |
| AACR GENIE V9.0 | 21 | 468 | 38 | 791 | 3 | 80 | 0 | 82 | 4 | 306 | 0 | 29 | 4 | 98 | 0 | 21 |
| Dubot et al., 2018 | 0 | 61 |  |  |  |  |  |  | 1 | 22 | 0 | 17 | 1 | 22 |  |  |
| Zehir et al., 2017 | 4 | 59 | 1 | 9 | 1 | 12 | 0 | 17 | 0 | 44 | 0 | 3 | 0 | 16 | 0 | 26 |
| Zehir et al., 2017 SG |  |  | 5 | 105 |  |  |  |  |  |  |  |  |  |  |  |  |
| Bersani et al., 2017 |  |  |  |  |  |  |  |  | 5 | 325 |  |  |  |  | 0 | 19 |
| Biswas et al., 2014 | 7 | 84 |  |  |  |  |  |  |  |  |  |  |  |  |  |  |
| Haft et al., 2019 |  |  |  |  |  |  |  |  | 2 | 46 |  |  |  |  |  |  |
| Jayaprakash et al., 2019 | 6 | 28 |  |  |  |  |  |  |  |  |  |  |  |  |  |  |
| Mirghani et al., 2018 |  |  |  |  |  |  |  |  | 0 | 62 |  |  |  |  |  |  |
| Nakagaki et al., 2017 | 2 | 47 |  |  |  |  |  |  |  |  |  |  |  |  |  |  |
| Ock et al., 2016 |  |  |  |  |  |  |  |  |  |  |  |  |  |  |  |  |
| Oikawa et al., 2016 | 6 | 220 |  |  |  |  |  |  |  |  |  |  |  |  |  |  |
| Smith et al., 2019 |  |  |  |  |  |  |  |  |  |  |  |  | 0 | 21 |  |  |
| Su et al., 2017 |  |  |  |  |  |  |  |  |  |  |  |  |  |  |  |  |
| Westbrook et al., 2019 |  |  |  |  |  |  |  |  |  |  |  |  |  |  |  |  |
| Samstein et al., 2019 | 2 | 56 |  |  | 1 | 6 | 0 | 14 | 2 | 31 | 0 | 8 | 0 | 14 | 0 | 9 |
| Robinson et al., 2017 | 0 | 8 |  |  | 0 | 1 |  |  | 1 | 2 |  |  | 0 | 1 | 1 | 2 |
| Robinson et al., 2017 |  |  | 1 | 14 |  |  |  |  |  |  |  |  |  |  |  |  |
| Priestley et al., 2019 | 0 | 12 | 1 | 19 | 0 | 10 | 0 | 1 | 0 | 3 | 0 | 5 | 1 | 2 | 0 | 1 |
| Popovic et al., 2010 | 13 | 60 |  |  |  |  |  |  |  |  |  |  |  |  | 8 | 39 |
| Pickering et al., 2014 |  |  |  |  |  |  |  |  |  |  |  |  |  |  |  |  |
| Leblanc et al., 2020 |  |  |  |  |  |  |  |  |  |  |  |  |  |  |  |  |
| Mueller et al., 2020 |  |  | 14 | 63 |  |  |  |  |  |  |  |  |  |  |  |  |
| Kim et al., 2020 |  |  | 4 | 42 |  |  |  |  |  |  |  |  |  |  |  |  |
| Hsieh et al., 2020 |  |  | 9 | 33 |  |  |  |  |  |  |  |  |  |  |  |  |
| Khoo et al., 2017 |  |  | 4 | 15 |  |  |  |  |  |  |  |  |  |  |  |  |
| Nakagaki et al., 2018 |  |  |  |  |  |  |  |  |  |  |  |  |  |  |  |  |
| Reder et al., 2021 |  |  |  |  |  |  |  |  | 9 | 56 |  |  |  |  |  |  |
| Patel et al., 2021 | 4 | 30 |  |  |  |  |  |  |  |  |  |  |  |  |  |  |
| Fonseca et al., 2015 |  |  | 0 | 17 |  |  |  |  |  |  |  |  |  |  |  |  |
| Li et al., 2018 |  |  |  |  |  |  | 34 | 168 |  |  |  |  |  |  |  |  |
| Lin et al., 2002 |  |  |  |  |  |  |  |  |  |  |  |  | 10 | 28 |  |  |
| Russo et al., 2006 |  |  |  |  |  |  |  |  |  |  |  |  | 5 | 81 |  |  |
| Kobyashi et al., 2019 | 6 | 153 |  |  |  |  |  |  | 1 | 32 | 0 | 74 | 0 | 25 |  |  |
| Dogan et al., 2019 |  |  | 6 | 25 |  |  |  |  |  |  |  |  |  |  |  |  |
| Ginkel et al., 2016 | 6 | 34 |  |  |  |  |  |  | 3 | 37 | 1 | 16 | 0 | 18 | 1 | 14 |
| Rampias et al., 2014 |  |  |  |  |  |  |  |  | 3 | 13 | 0 | 8 | 7 | 82 | 0 | 3 |

|  | **KRAS** | | | | | | | | | | | | | | | |
| --- | --- | --- | --- | --- | --- | --- | --- | --- | --- | --- | --- | --- | --- | --- | --- | --- |
|  | Oral cavity | | Salivary gland | | sinonasal | | nasopharynx | | oropharynx | | hypopharynx | | larynx | | other | |
| Cohorts | mutated | total | mutated | total | mutated | total | mutated | total | mutated | total | mutated | total | mutated | total | mutated | total |
| Das et al., 2000 | 4 | 12 |  |  |  |  |  |  |  |  |  |  |  |  |  |  |
| Yoo et al., 2000 |  |  | 2 | 24 |  |  |  |  |  |  |  |  |  |  |  |  |
| Yoo et al., 2000 |  |  |  |  |  |  |  |  |  |  |  |  |  |  |  |  |
| Weber et al., 2003 | 1 | 13 |  |  |  |  |  |  | 1 | 33 | 2 | 18 | 1 | 25 |  |  |
| Perrone et al., 2003 |  |  |  |  |  |  |  |  |  |  |  |  |  |  |  |  |
| Ruiz-Godoy et al., 2006 |  |  |  |  |  |  |  |  |  |  |  |  | 0 | 20 |  |  |
| Sathyan et al., 2006 |  |  |  |  |  |  |  |  |  |  |  |  |  |  |  |  |
| Frattini et al., 2006 |  |  |  |  | 9 | 18 |  |  |  |  |  |  |  |  |  |  |
| Augello et al., 2006 |  |  | 2 | 33 |  |  |  |  |  |  |  |  |  |  |  |  |
| Sathyan et al., 2007 | 0 | 152 |  |  |  |  |  |  |  |  |  |  |  |  |  |  |
| Gupta et al., 2007 | 0 | 20 |  |  |  |  | 0 | 1 |  |  |  |  | 0 | 14 |  |  |
| Sheikh Ali et al., 2008 | 0 | 51 |  |  |  |  |  |  | 0 | 9 | 0 | 14 | 0 | 17 |  |  |
| Chou et al., 2008 |  |  |  |  |  |  | 0 | 45 |  |  |  |  |  |  |  |  |
| Bornholdt et al., 2008 |  |  |  |  | 8 | 174 |  |  |  |  |  |  |  |  |  |  |
| Dahse et al., 2009 |  |  | 1 | 65 |  |  |  |  |  |  |  |  |  |  |  |  |
| Murugan et al., 2009 |  |  |  |  |  |  |  |  |  |  |  |  |  |  |  |  |
| Bruckman et al., 2010 | 1 | 42 |  |  |  |  |  |  |  |  |  |  |  |  |  |  |
| van Damme et al., 2010 |  |  |  |  |  |  |  |  | 1 | 22 |  |  |  |  |  |  |
| Chang et al., 2010 | 0 | 58 |  |  |  |  |  |  |  |  |  |  |  |  |  |  |
| Tetsu et al., 2010 | 1 | 8 | 0 | 3 | 0 | 1 |  |  |  |  |  |  |  |  | 0 | 5 |
| Murray et al., 2010 |  |  |  |  |  |  |  |  |  |  |  |  |  |  |  |  |
| Agarwal et al., 2011 | 0 | 75 |  |  |  |  |  |  | 0 | 21 | 0 | 9 | 0 | 13 |  |  |
| Stransky et al., 2011 | 0 | 51 |  |  | 0 | 2 |  |  | 0 | 15 | 1 | 9 | 0 | 15 |  |  |
| Cohen et al., 2011 | 0 | 37 |  |  |  |  |  |  |  |  |  |  |  |  |  |  |
| Trivedi et al., 2011 |  |  |  |  |  |  |  |  |  |  |  |  |  |  |  |  |
| Wang et al., 2011 | 0 | 47 |  |  |  |  |  |  |  |  |  |  |  |  |  |  |
| Friedland et al., 2011 |  |  |  |  |  |  |  |  | 0 | 20 |  |  |  |  |  |  |
| Szabo et al., 2011 |  |  |  |  |  |  |  |  |  |  |  |  |  |  |  |  |
| Szanyi et al., 2011 |  |  |  |  |  |  |  |  |  |  |  |  |  |  |  |  |
| Suda et al., 2012 | 2 | 31 |  |  | 0 | 11 |  |  | 1 | 25 | 0 | 25 | 0 | 23 |  |  |
| Koumaki et al., 2012 |  |  |  |  |  |  |  |  |  |  |  |  |  |  |  |  |
| Lopez et al., 2012 and Garcia-Inclan et al., 2012 |  |  |  |  | 7 | 58 |  |  |  |  |  |  |  |  |  |  |
| Smilek et al., 2012 |  |  |  |  |  |  |  |  |  |  |  |  |  |  |  |  |
| Bissada et al., 2013 | 0 | 21 |  |  |  |  |  |  | 4 | 123 | 0 | 11 | 3 | 29 | 0 | 11 |
| Choisea et al., 2013 |  |  |  |  |  |  |  |  |  |  |  |  |  |  |  |  |
| Lechner et al., 2013 |  |  |  |  | ` |  |  |  | 2 | 34 |  |  |  |  |  |  |
| Pickering et al., 2013 | 0 | 40 |  |  |  |  |  |  |  |  |  |  |  |  |  |  |
| Ho et al., 2013 |  |  | 0 | 59 |  |  |  |  |  |  |  |  |  |  |  |  |
| Stephens et al., 2013 |  |  | 0 | 24 |  |  |  |  |  |  |  |  |  |  |  |  |
| Liu et al., 2013 |  |  |  |  |  |  |  |  |  |  |  |  |  |  |  |  |
| Cros et al., 2013 |  |  | 4 | 107 |  |  |  |  |  |  |  |  |  |  |  |  |
| Fuji et al., 2013 |  |  |  |  |  |  |  |  | 0 | 71 | 0 | 83 | 0 | 51 |  |  |
| Projetti et al., 2013 |  |  |  |  | 2 | 34 |  |  |  |  |  |  |  |  |  |  |
| Szablewski et al., 2013 |  |  |  |  | 12 | 28 |  |  |  |  |  |  |  |  |  |  |
| Carvalho et al., 2013 | 0 | 66 |  |  |  |  |  |  | 0 | 12 | 0 | 2 | 0 | 14 |  |  |
| Zanaruddin et al., 2013 | 0 | 107 |  |  |  |  |  |  |  |  |  |  |  |  |  |  |
| Wetterskog et al., 2013 |  |  |  |  |  |  |  |  |  |  |  |  |  |  |  |  |
| Fury et al., 2013 | 1 | 1 |  |  | 0 | 1 | 0 | 1 | 0 | 14 | 0 | 1 |  |  |  |  |
| Chang et al., 2014 | 0 | 79 |  |  |  |  |  |  |  |  |  |  |  |  |  |  |
| Zhang et al., 2014 |  |  |  |  |  |  |  |  |  |  |  |  |  |  |  |  |
| Lin et al., 2014 |  |  |  |  |  |  | 1 | 56 |  |  |  |  |  |  |  |  |
| Ross et al., 2014 |  |  | 0 | 15 |  |  |  |  |  |  |  |  |  |  |  |  |
| Al Rawi et al., 2014 |  |  |  |  |  |  |  |  |  |  |  |  |  |  |  |  |
| Prigge et al., 2014 | 0 | 3 |  |  |  |  |  |  | 0 | 21 | 0 | 1 |  |  |  |  |
| Tan et al., 2014 | 0 | 66 |  |  |  |  |  |  |  |  |  |  |  |  |  |  |
| Chung et al., 2014 |  |  |  |  |  |  |  |  |  |  |  |  |  |  |  |  |
| Boeckx et al., 2014 |  |  |  |  |  |  |  |  |  |  |  |  |  |  |  |  |
| Choisea et al., 2014 |  |  |  |  |  |  |  |  |  |  |  |  |  |  |  |  |
| Lin et al., 2014 |  |  |  |  |  |  | 1 | 66 |  |  |  |  |  |  |  |  |
| Zhang et al., 2014 |  |  |  |  |  |  | 0 | 123 |  |  |  |  |  |  |  |  |
| Franchi et al., 2014 |  |  |  |  | 1 | 27 |  |  |  |  |  |  |  |  |  |  |
| Zhang et al., 2015 |  |  |  |  |  |  | 1 | 70 |  |  |  |  |  |  |  |  |
| Seiwert et al., 2015 |  |  |  |  |  |  |  |  |  |  |  |  |  |  |  |  |
| Vettore et al., 2015 |  |  |  |  |  |  |  |  |  |  |  |  |  |  |  |  |
| Chen et al., 2015 | 8 | 345 |  |  |  |  |  |  |  |  |  |  |  |  |  |  |
| Fu et al., 2015 |  |  | 2 | 18 |  |  |  |  |  |  |  |  |  |  |  |  |
| Grunewald et al., 2015 |  |  | 0 | 84 |  |  |  |  |  |  |  |  |  |  |  |  |
| Kato et al., 2015 |  |  |  |  |  |  |  |  |  |  |  |  |  |  |  |  |
| Choisea et al., 2015 |  |  | 0 | 29 |  |  |  |  |  |  |  |  |  |  |  |  |
| Braig et al., 2016 | 0 | 12 |  |  | 0 | 2 |  |  | 0 | 19 | 1 | 9 | 1 | 4 |  |  |
| Rettig et al., 2016 |  |  | 0 | 25 |  |  |  |  |  |  |  |  |  |  |  |  |
| Mitani et al., 2016 |  |  | 0 | 65 |  |  |  |  |  |  |  |  |  |  |  |  |
| Drier et al., 2016 |  |  | 0 | 10 |  |  |  |  |  |  |  |  |  |  |  |  |
| Shalmon et al., 2016 |  |  | 0 | 21 |  |  |  |  |  |  |  |  |  |  |  |  |
| Wang et al., 2016 |  |  | 0 | 149 |  |  |  |  |  |  |  |  |  |  |  |  |
| Tinhofer et al., 2016 |  |  |  |  |  |  |  |  |  |  |  |  |  |  |  |  |
| Kucuk et al., 2016 | 0 | 26 |  |  |  |  |  |  |  |  |  |  |  |  |  |  |
| Schneider et al., 2016 |  |  | 0 | 43 |  |  |  |  |  |  |  |  |  |  |  |  |
| Choisea et al., 2016 |  |  |  |  |  |  |  |  |  |  |  |  |  |  |  |  |
| Dalin et al., 2016 |  |  | 0 | 31 |  |  |  |  |  |  |  |  |  |  |  |  |
| Udager et al., 2016 |  |  |  |  | 56 | 56 |  |  |  |  |  |  |  |  |  |  |
| Wu et al., 2016 | 1 | 56 |  |  |  |  |  |  | 8 | 76 |  |  | 9 | 82 |  |  |
| Chau et al., 2016 | 1 | 60 |  |  | 0 | 5 | 0 | 7 | 0 | 97 | 0 | 6 | 0 | 24 | 0 | 14 |
| Bell et al., 2016 |  |  |  |  |  |  |  |  |  |  |  |  |  |  | 10 | 24 |
| Luk et al., 2016 |  |  | 0 | 23 |  |  |  |  |  |  |  |  |  |  |  |  |
| Al-Hebshi et al., 2016 |  |  |  |  |  |  |  |  |  |  |  |  |  |  |  |  |
| Hedberg et al., 2016 | 0 | 6 |  |  |  |  |  |  |  |  | 0 | 3 | 1 | 4 |  |  |
| Chuerduangphui et al., 2017 |  |  |  |  |  |  |  |  |  |  |  |  |  |  |  |  |
| Morris et al., 2017 | 1 | 26 | 4 | 32 | 1 | 15 | 0 | 9 | 0 | 23 | 0 | 2 | 0 | 8 | 3 | 36 |
| Yue et al., 2017 |  |  |  |  |  |  |  |  |  |  |  |  |  |  |  |  |
| Abdolkarim Moazeni-Roodi et al., 2017 |  |  |  |  |  |  |  |  |  |  |  |  |  |  |  |  |
| Zhang et al., 2017 |  |  |  |  |  |  | 3 | 94 |  |  |  |  |  |  |  |  |
| Ali et al., 2017 |  |  |  |  |  |  |  |  |  |  |  |  |  |  |  |  |
| Upadhyay et al., 2017 | 0 | 25 |  |  |  |  |  |  |  |  |  |  |  |  |  |  |
| kang et al., 2017 |  |  | 0 | 18 |  |  |  |  |  |  |  |  |  |  |  |  |
| Dalin et al., 2017 |  |  | 0 | 40 |  |  |  |  |  |  |  |  |  |  |  |  |
| Li et al., 2017 |  |  |  |  |  |  | 1 | 105 |  |  |  |  |  |  |  |  |
| Dogan et al., 2017 |  |  |  |  | 3 | 30 |  |  |  |  |  |  |  |  |  |  |
| Krishna et al., 2018 |  |  |  |  |  |  |  |  |  |  |  |  |  |  |  |  |
| Lin et al., 2018 |  |  |  |  |  |  |  |  |  |  |  |  |  |  |  |  |
| Saida et al., 2018 |  |  | 6 | 70 |  |  |  |  |  |  |  |  |  |  |  |  |
| Hallani et al., 2018 |  |  | 0 | 23 |  |  |  |  |  |  |  |  |  |  |  |  |
| Shimura et al., 2018 |  |  | 0 | 140 |  |  |  |  |  |  |  |  |  |  |  |  |
| Perdomo et al., 2018 |  |  |  |  |  |  |  |  |  |  |  |  |  |  |  |  |
| Vossen etal., 2018 |  |  |  |  |  |  |  |  |  |  |  |  |  |  |  |  |
| Batta et al., 2019 | 0 | 39 |  |  |  |  |  |  |  |  |  |  |  |  | 0 | 7 |
| Akagi et al., 2019 |  |  |  |  |  |  |  |  |  |  |  |  |  |  |  |  |
| Chung et al., 2019 |  |  |  |  |  |  |  |  |  |  |  |  |  |  |  |  |
| Reder et al., 2019 |  |  |  |  |  |  |  |  | 0 | 12 |  |  |  |  |  |  |
| Stanek et al., 2019 |  |  |  |  |  |  |  |  |  |  |  |  |  |  |  |  |
| Urano et al., 2019 |  |  |  |  |  |  |  |  |  |  |  |  |  |  |  |  |
| Nakaguro et al., 2019 |  |  |  |  |  |  |  |  |  |  |  |  |  |  |  |  |
| Wang et al., 2019 |  |  |  |  | 30 | 80 |  |  |  |  |  |  |  |  |  |  |
| Reder et al., 2019 |  |  |  |  |  |  |  |  | 1 | 12 |  |  |  |  |  |  |
| Gauthaman et al., 2020 | 25 | 44 |  |  |  |  |  |  | 2 | 3 | 4 | 6 | 0 | 0 | 3 | 4 |
| Kyurkchiyan et al., 2020 |  |  |  |  |  |  |  |  |  |  |  |  | 2 | 57 |  |  |
| ORCA ICGC |  |  |  |  |  |  |  |  |  |  |  |  |  |  |  |  |
| Sasaki et al., 2020 | 1 | 12 |  |  |  |  |  |  |  |  | 11 | 20 | 2 | 7 | 4 | 12 |
| Morita et al., 2020 |  |  | 7 | 101 |  |  |  |  |  |  |  |  |  |  |  |  |
| Morfouace et al., 2020 |  |  |  |  | 0 | 3 | 0 | 10 |  |  |  |  |  |  | 0 | 1 |
| Kawamura et al., 2020 | 0 | 20 |  |  |  |  |  |  |  |  |  |  |  |  |  |  |
| Pérez Sayáns et al 2019 | 1 | 303 |  |  |  |  |  |  | 0 | 82 | 0 | 10 | 0 | 117 |  |  |
| Masato et al., 2021 |  |  |  |  |  |  |  |  |  |  |  |  |  |  |  |  |
| Sanchez-Fernandez et al., 2021 |  |  |  |  |  |  |  |  |  |  |  |  |  |  | 7 | 48 |
| AACR GENIE V9.0 | 8 | 468 | 8 | 791 | 0 | 80 | 2 | 82 | 4 | 306 | 0 | 29 | 4 | 98 | 0 | 21 |
| Dubot et al., 2018 | 0 | 61 |  |  |  |  |  |  | 2 | 22 | 0 | 17 | 0 | 22 |  |  |
| Zehir et al., 2017 | 1 | 59 | 0 | 9 | 0 | 12 | 0 | 17 | 0 | 52 | 0 | 3 | 1 | 16 | 0 | 18 |
| Zehir et al., 2017 SG |  |  | 1 | 105 |  |  |  |  |  |  |  |  |  |  |  |  |
| Bersani et al., 2017 |  |  |  |  |  |  |  |  | 9 | 325 |  |  |  |  | 0 | 19 |
| Biswas et al., 2014 |  |  |  |  |  |  |  |  |  |  |  |  |  |  |  |  |
| Haft et al., 2019 |  |  |  |  |  |  |  |  | 0 | 46 |  |  |  |  |  |  |
| Jayaprakash et al., 2019 | 0 | 28 |  |  |  |  |  |  |  |  |  |  |  |  |  |  |
| Mirghani et al., 2018 |  |  |  |  |  |  |  |  | 2 | 62 |  |  |  |  |  |  |
| Nakagaki et al., 2017 | 0 | 47 |  |  |  |  |  |  |  |  |  |  |  |  |  |  |
| Ock et al., 2016 |  |  |  |  |  |  |  |  |  |  |  |  |  |  |  |  |
| Oikawa et al., 2016 | 1 | 220 |  |  |  |  |  |  |  |  |  |  |  |  |  |  |
| Smith et al., 2019 |  |  |  |  |  |  |  |  |  |  |  |  |  |  |  |  |
| Su et al., 2017 |  |  |  |  |  |  |  |  |  |  |  |  | 1 | 21 |  |  |
| Westbrook et al., 2019 | 0 | 6 |  |  |  |  | 0 | 1 | 0 | 7 | 0 | 2 | 1 | 7 |  |  |
| Samstein et al., 2019 | 1 | 56 |  |  | 0 | 6 | 0 | 14 | 1 | 33 | 0 | 8 | 1 | 14 | 0 | 9 |
| Robinson et al., 2017 | 0 | 8 |  |  | 0 | 1 |  |  | 0 | 2 |  |  | 0 | 1 | 1 | 2 |
| Robinson et al., 2017 |  |  | 0 | 14 |  |  |  |  |  |  |  |  |  |  |  |  |
| Priestley et al., 2019 | 0 | 12 | 0 | 19 | 0 | 10 | 0 | 1 | 0 | 3 | 0 | 5 | 0 | 2 | 0 | 1 |
| Popovic et al., 2010 |  |  |  |  |  |  |  |  |  |  |  |  |  |  | 0 | 39 |
| Pickering et al., 2014 |  |  |  |  |  |  |  |  |  |  |  |  |  |  |  |  |
| Leblanc et al., 2020 |  |  |  |  |  |  |  |  |  |  |  |  |  |  |  |  |
| Mueller et al., 2020 |  |  | 3 | 63 |  |  |  |  |  |  |  |  |  |  |  |  |
| Kim et al., 2020 |  |  | 0 | 42 |  |  |  |  |  |  |  |  |  |  |  |  |
| Hsieh et al., 2020 |  |  |  |  |  |  |  |  |  |  |  |  |  |  |  |  |
| Khoo et al., 2017 |  |  | 4 | 15 |  |  |  |  |  |  |  |  |  |  |  |  |
| Nakagaki et al., 2018 |  |  |  |  |  |  |  |  |  |  |  |  |  |  |  |  |
| Reder et al., 2021 |  |  |  |  |  |  |  |  | 8 | 56 |  |  |  |  |  |  |
| Patel et al., 2021 | 0 | 30 |  |  |  |  |  |  |  |  |  |  |  |  |  |  |
| Fonseca et al., 2015 |  |  | 3 | 17 |  |  |  |  |  |  |  |  |  |  |  |  |
| Li et al., 2018 |  |  |  |  |  |  |  |  |  |  |  |  |  |  |  |  |
| Lin et al., 2002 |  |  |  |  |  |  |  |  |  |  |  |  |  |  |  |  |
| Russo et al., 2006 |  |  |  |  |  |  |  |  |  |  |  |  | 0 | 81 |  |  |
| Kobyashi et al., 2019 |  |  |  |  |  |  |  |  |  |  |  |  |  |  |  |  |
| Dogan et al., 2019 |  |  | 0 | 25 |  |  |  |  |  |  |  |  |  |  |  |  |
| Ginkel et al., 2016 |  |  |  |  |  |  |  |  | 1 | 37 |  |  |  |  | 1 | 14 |
| Rampias et al., 2014 |  |  |  |  |  |  |  |  |  |  |  |  |  |  |  |  |

|  | **NRAS** | | | | | | | | | | | | | | | |
| --- | --- | --- | --- | --- | --- | --- | --- | --- | --- | --- | --- | --- | --- | --- | --- | --- |
|  | Oral cavity | | Salivary gland | | sinonasal | | nasopharynx | | oropharinx | | hypopharynx | | larynx | | Other | |
| Cohorts | mutated | total | mutated | total | mutated | total | mutated | total | mutated | total | mutated | total | mutated | total | mutated | total |
| Das et al., 2000 |  |  |  |  |  |  |  |  |  |  |  |  |  |  |  |  |
| Yoo et al., 2000 |  |  |  |  |  |  |  |  |  |  |  |  |  |  |  |  |
| Yoo et al., 2000 |  |  |  |  |  |  |  |  |  |  |  |  |  |  |  |  |
| Weber et al., 2003 |  |  |  |  |  |  |  |  |  |  |  |  |  |  |  |  |
| Perrone et al., 2003 |  |  |  |  |  |  |  |  |  |  |  |  |  |  |  |  |
| Ruiz-Godoy et al., 2006 |  |  |  |  |  |  |  |  |  |  |  |  |  |  |  |  |
| Sathyan et al., 2006 |  |  |  |  |  |  |  |  |  |  |  |  |  |  |  |  |
| Frattini et al., 2006 |  |  |  |  |  |  |  |  |  |  |  |  |  |  |  |  |
| Augello et al., 2006 |  |  |  |  |  |  |  |  |  |  |  |  |  |  |  |  |
| Sathyan et al., 2007 | 0 | 152 |  |  |  |  |  |  |  |  |  |  |  |  |  |  |
| Gupta et al., 2007 | 0 | 20 |  |  |  |  | 0 | 1 |  |  |  |  | 0 | 14 |  |  |
| Sheikh Ali et al., 2008 |  |  |  |  |  |  |  |  |  |  |  |  |  |  |  |  |
| Chou et al., 2008 |  |  |  |  |  |  |  |  |  |  |  |  |  |  |  |  |
| Bornholdt et al., 2008 |  |  |  |  |  |  |  |  |  |  |  |  |  |  |  |  |
| Dahse et al., 2009 |  |  |  |  |  |  |  |  |  |  |  |  |  |  |  |  |
| Murugan et al., 2009 |  |  |  |  |  |  |  |  |  |  |  |  |  |  |  |  |
| Bruckman et al., 2010 |  |  |  |  |  |  |  |  |  |  |  |  |  |  |  |  |
| van Damme et al., 2010 |  |  |  |  |  |  |  |  |  |  |  |  |  |  |  |  |
| Chang et al., 2010 | 0 | 58 |  |  |  |  |  |  |  |  |  |  |  |  |  |  |
| Tetsu et al., 2010 | 0 | 8 | 1 | 3 | 0 | 1 |  |  |  |  |  |  |  |  | 0 | 5 |
| Murray et al., 2010 |  |  |  |  |  |  |  |  |  |  |  |  |  |  |  |  |
| Agarwal et al., 2011 | 0 | 75 |  |  |  |  |  |  | 0 | 21 | 0 | 9 | 0 | 13 |  |  |
| Stransky et al., 2011 | 0 | 51 |  |  | 0 | 2 |  |  | 0 | 15 | 0 | 9 | 0 | 15 |  |  |
| Cohen et al., 2011 | 0 | 37 |  |  |  |  |  |  |  |  |  |  |  |  |  |  |
| Trivedi et al., 2011 |  |  |  |  |  |  |  |  |  |  |  |  |  |  |  |  |
| Wang et al., 2011 |  |  |  |  |  |  |  |  |  |  |  |  |  |  |  |  |
| Friedland et al., 2011 |  |  |  |  |  |  |  |  |  |  |  |  |  |  |  |  |
| Szabo et al., 2011 |  |  |  |  |  |  |  |  |  |  |  |  |  |  |  |  |
| Szanyi et al., 2011 |  |  |  |  |  |  |  |  |  |  |  |  |  |  |  |  |
| Suda et al., 2012 |  |  |  |  |  |  |  |  |  |  |  |  |  |  |  |  |
| Koumaki et al., 2012 |  |  |  |  |  |  |  |  |  |  |  |  |  |  |  |  |
| Lopez et al., 2012 and Garcia-Inclan et al., 2012 |  |  |  |  |  |  |  |  |  |  |  |  |  |  |  |  |
| Smilek et al., 2012 |  |  |  |  |  |  |  |  |  |  |  |  |  |  |  |  |
| Bissada et al., 2013 |  |  |  |  |  |  |  |  |  |  |  |  |  |  |  |  |
| Choisea et al., 2013 |  |  |  |  |  |  |  |  |  |  |  |  |  |  |  |  |
| Lechner et al., 2013 |  |  |  |  |  |  |  |  | 0 | 34 |  |  |  |  |  |  |
| Pickering et al., 2013 | 0 | 40 |  |  |  |  |  |  |  |  |  |  |  |  |  |  |
| Ho et al., 2013 |  |  | 1 | 59 |  |  |  |  |  |  |  |  |  |  |  |  |
| Stephens et al., 2013 |  |  | 0 | 24 |  |  |  |  |  |  |  |  |  |  |  |  |
| Liu et al., 2013 |  |  |  |  |  |  |  |  |  |  |  |  |  |  |  |  |
| Cros et al., 2013 |  |  | 2 | 107 |  |  |  |  |  |  |  |  |  |  |  |  |
| Fuji et al., 2013 |  |  |  |  |  |  |  |  |  |  |  |  |  |  |  |  |
| Projetti et al., 2013 |  |  |  |  |  |  |  |  |  |  |  |  |  |  |  |  |
| Szablewski et al., 2013 |  |  |  |  |  |  |  |  |  |  |  |  |  |  |  |  |
| Carvalho et al., 2013 |  |  |  |  |  |  |  |  |  |  |  |  |  |  |  |  |
| Zanaruddin et al., 2013 | 0 | 107 |  |  |  |  |  |  |  |  |  |  |  |  |  |  |
| Wetterskog et al., 2013 |  |  |  |  |  |  |  |  |  |  |  |  |  |  |  |  |
| Fury et al., 2013 | 1 | 1 |  |  | 0 | 1 | 0 | 1 | 0 | 14 | 0 | 1 |  |  |  |  |
| Chang et al., 2014 | 0 | 79 |  |  |  |  |  |  |  |  |  |  |  |  |  |  |
| Zhang et al., 2014 |  |  |  |  |  |  |  |  |  |  |  |  |  |  |  |  |
| Lin et al., 2014 |  |  |  |  |  |  | 1 | 56 |  |  |  |  |  |  |  |  |
| Ross et al., 2014 |  |  | 0 | 15 |  |  |  |  |  |  |  |  |  |  |  |  |
| Al Rawi et al., 2014 |  |  |  |  |  |  |  |  |  |  |  |  |  |  |  |  |
| Prigge et al., 2014 |  |  |  |  |  |  |  |  |  |  |  |  |  |  |  |  |
| Tan et al., 2014 |  |  |  |  |  |  |  |  |  |  |  |  |  |  |  |  |
| Chung et al., 2014 |  |  |  |  |  |  |  |  |  |  |  |  |  |  |  |  |
| Boeckx et al., 2014 |  |  |  |  |  |  |  |  |  |  |  |  |  |  |  |  |
| Choisea et al., 2014 |  |  |  |  |  |  |  |  |  |  |  |  |  |  |  |  |
| Lin et al., 2014 |  |  |  |  |  |  | 0 | 66 |  |  |  |  |  |  |  |  |
| Zhang et al., 2014 |  |  |  |  |  |  | 5 | 123 |  |  |  |  |  |  |  |  |
| Franchi et al., 2014 |  |  |  |  |  |  |  |  |  |  |  |  |  |  |  |  |
| Zhang et al., 2015 |  |  |  |  |  |  |  |  |  |  |  |  |  |  |  |  |
| Seiwert et al., 2015 |  |  |  |  |  |  |  |  |  |  |  |  |  |  |  |  |
| Vettore et al., 2015 |  |  |  |  |  |  |  |  |  |  |  |  |  |  |  |  |
| Chen et al., 2015 |  |  |  |  |  |  |  |  |  |  |  |  |  |  |  |  |
| Fu et al., 2015 |  |  | 0 | 18 |  |  |  |  |  |  |  |  |  |  |  |  |
| Grunewald et al., 2015 |  |  | 2 | 84 |  |  |  |  |  |  |  |  |  |  |  |  |
| Kato et al., 2015 |  |  |  |  |  |  |  |  |  |  |  |  |  |  |  |  |
| Choisea et al., 2015 |  |  | 0 | 29 |  |  |  |  |  |  |  |  |  |  |  |  |
| Braig et al., 2016 | 1 | 12 |  |  | 0 | 2 |  |  | 0 | 19 | 0 | 9 | 1 | 4 |  |  |
| Rettig et al., 2016 |  |  | 1 | 25 |  |  |  |  |  |  |  |  |  |  |  |  |
| Mitani et al., 2016 |  |  | 0 | 65 |  |  |  |  |  |  |  |  |  |  |  |  |
| Drier et al., 2016 |  |  | 0 | 10 |  |  |  |  |  |  |  |  |  |  |  |  |
| Shalmon et al., 2016 |  |  |  |  |  |  |  |  |  |  |  |  |  |  |  |  |
| Wang et al., 2016 |  |  | 0 | 149 |  |  |  |  |  |  |  |  |  |  |  |  |
| Tinhofer et al., 2016 |  |  |  |  |  |  |  |  |  |  |  |  |  |  |  |  |
| Kucuk et al., 2016 |  |  |  |  |  |  |  |  |  |  |  |  |  |  |  |  |
| Schneider et al., 2016 |  |  |  |  |  |  |  |  |  |  |  |  |  |  |  |  |
| Choisea et al., 2016 |  |  |  |  |  |  |  |  |  |  |  |  |  |  |  |  |
| Dalin et al., 2016 |  |  | 0 | 31 |  |  |  |  |  |  |  |  |  |  |  |  |
| Udager et al., 2016 |  |  |  |  |  |  |  |  |  |  |  |  |  |  |  |  |
| Wu et al., 2016 |  |  |  |  |  |  |  |  |  |  |  |  |  |  |  |  |
| Chau et al., 2016 | 2 | 60 |  |  | 0 | 5 | 0 | 7 | 0 | 97 | 0 | 6 | 0 | 24 | 0 | 14 |
| Bell et al., 2016 |  |  |  |  |  |  |  |  |  |  |  |  |  |  | 2 | 24 |
| Luk et al., 2016 |  |  | 1 | 23 |  |  |  |  |  |  |  |  |  |  |  |  |
| Al-Hebshi et al., 2016 |  |  |  |  |  |  |  |  |  |  |  |  |  |  |  |  |
| Hedberg et al., 2016 | 1 | 6 |  |  |  |  |  |  |  |  | 0 | 3 | 0 | 4 |  |  |
| Chuerduangphui et al., 2017 |  |  |  |  |  |  |  |  |  |  |  |  |  |  |  |  |
| Morris et al., 2017 | 1 | 26 | 0 | 32 | 0 | 15 | 1 | 9 | 0 | 23 | 0 | 2 | 0 | 8 | 0 | 36 |
| Yue et al., 2017 |  |  |  |  |  |  |  |  |  |  |  |  |  |  |  |  |
| Abdolkarim Moazeni-Roodi et al., 2017 |  |  |  |  |  |  |  |  |  |  |  |  |  |  |  |  |
| Zhang et al., 2017 |  |  |  |  |  |  | 1 | 94 |  |  |  |  |  |  |  |  |
| Ali et al., 2017 |  |  |  |  |  |  |  |  |  |  |  |  |  |  |  |  |
| Upadhyay et al., 2017 | 0 | 24 |  |  |  |  |  |  |  |  |  |  |  |  |  |  |
| kang et al., 2017 |  |  | 0 | 18 |  |  |  |  |  |  |  |  |  |  |  |  |
| Dalin et al., 2017 |  |  | 0 | 40 |  |  |  |  |  |  |  |  |  |  |  |  |
| Li et al., 2017 |  |  |  |  |  |  | 3 | 105 |  |  |  |  |  |  |  |  |
| Dogan et al., 2017 |  |  |  |  | 0 | 30 |  |  |  |  |  |  |  |  |  |  |
| Krishna et al., 2018 |  |  |  |  |  |  |  |  |  |  |  |  |  |  |  |  |
| Lin et al., 2018 |  |  |  |  |  |  |  |  |  |  |  |  |  |  |  |  |
| Saida et al., 2018 |  |  | 0 | 70 |  |  |  |  |  |  |  |  |  |  |  |  |
| Perdomo et al., 2018 |  |  |  |  |  |  |  |  |  |  |  |  |  |  |  |  |
| Vossen etal., 2018 |  |  |  |  |  |  |  |  |  |  |  |  |  |  |  |  |
| Hallani et al., 2018 |  |  | 0 | 23 |  |  |  |  |  |  |  |  |  |  |  |  |
| Shimura et al., 2018 |  |  | 0 | 140 |  |  |  |  |  |  |  |  |  |  |  |  |
| Batta et al., 2019 | 0 | 39 |  |  |  |  |  |  |  |  |  |  |  |  | 0 | 7 |
| Akagi et al., 2019 |  |  |  |  |  |  |  |  |  |  |  |  |  |  |  |  |
| Chung et al., 2019 |  |  |  |  |  |  |  |  |  |  |  |  |  |  |  |  |
| Reder et al., 2019 |  |  |  |  |  |  |  |  |  |  |  |  |  |  |  |  |
| Stanek et al., 2019 |  |  |  |  |  |  |  |  | 1 | 12 |  |  |  |  |  |  |
| Urano et al., 2019 |  |  |  |  |  |  |  |  |  |  |  |  |  |  |  |  |
| Nakaguro et al., 2019 |  |  |  |  |  |  |  |  |  |  |  |  |  |  |  |  |
| Wang et al., 2019 |  |  |  |  |  |  |  |  |  |  |  |  |  |  |  |  |
| Reder et al., 2019 |  |  |  |  |  |  |  |  | 0 | 12 |  |  |  |  |  |  |
| Gauthaman et al., 2020 |  |  |  |  |  |  |  |  |  |  |  |  |  |  |  |  |
| Kyurkchiyan et al., 2020 |  |  |  |  |  |  |  |  |  |  |  |  | 1 | 57 |  |  |
| ORCA ICGC |  |  |  |  |  |  |  |  |  |  |  |  |  |  |  |  |
| Sasaki et al., 2020 |  |  |  |  |  |  |  |  |  |  |  |  |  |  |  |  |
| Morita et al., 2020 |  |  | 0 | 101 |  |  |  |  |  |  |  |  |  |  |  |  |
| Morfouace et al., 2020 |  |  |  |  | 0 | 3 | 0 | 10 |  |  |  |  |  |  | 0 | 1 |
| Kawamura et al., 2020 | 0 | 20 |  |  |  |  |  |  |  |  |  |  |  |  |  |  |
| Pérez Sayáns et al 2019 | 0 | 303 |  |  |  |  |  |  | 0 | 82 | 0 | 10 | 1 | 117 |  |  |
| Masato et al., 2021 |  |  |  |  |  |  |  |  |  |  |  |  |  |  |  |  |
| Sanchez-Fernandez et al., 2021 |  |  |  |  |  |  |  |  |  |  |  |  |  |  | 2 | 48 |
| AACR GENIE V9.0 | 5 | 468 | 5 | 791 | 1 | 80 | 1 | 82 | 3 | 306 | 0 | 29 | 0 | 98 | 0 | 21 |
| Dubot et al., 2018 | 1 | 61 |  |  |  |  |  |  | 0 | 22 | 0 | 17 | 0 | 22 |  |  |
| Zehir et al., 2017 | 1 | 59 | 0 | 9 | 0 | 12 | 1 | 17 | 0 | 52 | 0 | 3 | 0 | 16 | 1 | 18 |
| Zehir et al., 2017 SG |  |  | 1 | 105 |  |  |  |  |  |  |  |  |  |  |  |  |
| Bersani et al., 2017 |  |  |  |  |  |  |  |  | 4 | 325 |  |  |  |  | 0 | 19 |
| Biswas et al., 2014 |  |  |  |  |  |  |  |  |  |  |  |  |  |  |  |  |
| Haft et al., 2019 |  |  |  |  |  |  |  |  | 0 | 46 |  |  |  |  |  |  |
| Jayaprakash et al., 2019 | 0 | 28 |  |  |  |  |  |  |  |  |  |  |  |  |  |  |
| Mirghani et al., 2018 |  |  |  |  |  |  |  |  | 0 | 62 |  |  |  |  |  |  |
| Nakagaki et al., 2017 | 0 | 47 |  |  |  |  |  |  |  |  |  |  |  |  |  |  |
| Ock et al., 2016 |  |  |  |  |  |  |  |  |  |  |  |  |  |  |  |  |
| Oikawa et al., 2016 | 2 | 220 |  |  |  |  |  |  |  |  |  |  |  |  |  |  |
| Smith et al., 2019 |  |  |  |  |  |  |  |  |  |  |  |  | 0 | 21 |  |  |
| Su et al., 2017 |  |  |  |  |  |  |  |  |  |  |  |  |  |  |  |  |
| Westbrook et al., 2019 | 0 | 6 |  |  |  |  | 0 | 1 | 0 | 7 | 0 | 2 | 0 | 7 |  |  |
| Samstein et al., 2019 | 1 | 56 |  |  | 0 | 6 | 1 | 14 | 1 | 33 | 0 | 8 | 0 | 14 | 0 | 9 |
| Robinson et al., 2017 | 0 | 8 |  |  | 0 | 1 |  |  | 0 | 2 |  |  | 0 | 1 | 0 | 2 |
| Robinson et al., 2017 |  |  | 0 | 14 |  |  |  |  |  |  |  |  |  |  |  |  |
| Priestley et al., 2019 | 0 | 12 | 0 | 19 | 0 | 10 | 0 | 1 | 0 | 3 | 0 | 5 | 0 | 2 | 0 | 1 |
| Popovic et al., 2010 |  |  |  |  |  |  |  |  |  |  |  |  |  |  | 2 | 39 |
| Pickering et al., 2014 |  |  |  |  |  |  |  |  |  |  |  |  |  |  |  |  |
| Leblanc et al., 2020 |  |  |  |  |  |  |  |  |  |  |  |  |  |  |  |  |
| Mueller et al., 2020 |  |  | 0 | 63 |  |  |  |  |  |  |  |  |  |  |  |  |
| Kim et al., 2020 |  |  | 0 | 42 |  |  |  |  |  |  |  |  |  |  |  |  |
| Hsieh et al., 2020 |  |  | 8 | 33 |  |  |  |  |  |  |  |  |  |  |  |  |
| Khoo et al., 2017 |  |  | 4 | 15 |  |  |  |  |  |  |  |  |  |  |  |  |
| Nakagaki et al., 2018 |  |  |  |  |  |  |  |  |  |  |  |  |  |  |  |  |
| Reder et al., 2021 |  |  |  |  |  |  |  |  | 7 | 56 |  |  |  |  |  |  |
| Patel et al., 2021 | 0 | 30 |  |  |  |  |  |  |  |  |  |  |  |  |  |  |
| Fonseca et al., 2015 |  |  | 1 | 17 |  |  |  |  |  |  |  |  |  |  |  |  |
| Li et al., 2018 |  |  |  |  |  |  |  |  |  |  |  |  |  |  |  |  |
| Lin et al., 2002 |  |  |  |  |  |  |  |  |  |  |  |  |  |  |  |  |
| Russo et al., 2006 |  |  |  |  |  |  |  |  |  |  |  |  | 0 | 81 |  |  |
| Kobyashi et al., 2019 |  |  |  |  |  |  |  |  |  |  |  |  |  |  |  |  |
| Dogan et al., 2019 |  |  | 0 | 25 |  |  |  |  |  |  |  |  |  |  |  |  |
| Ginkel et al., 2016 |  |  |  |  |  |  |  |  |  |  |  |  |  |  |  |  |
| Rampias et al., 2014 |  |  |  |  |  |  |  |  |  |  |  |  |  |  |  |  |

**eFigure 2: Forest plot of RAS mutation frequency according to geographical region**

Forest plot of RAS mutation frequency [%] in head and neck cancer according to geographical region. CI: Confidence interval. I^2^: Inconsistency index.

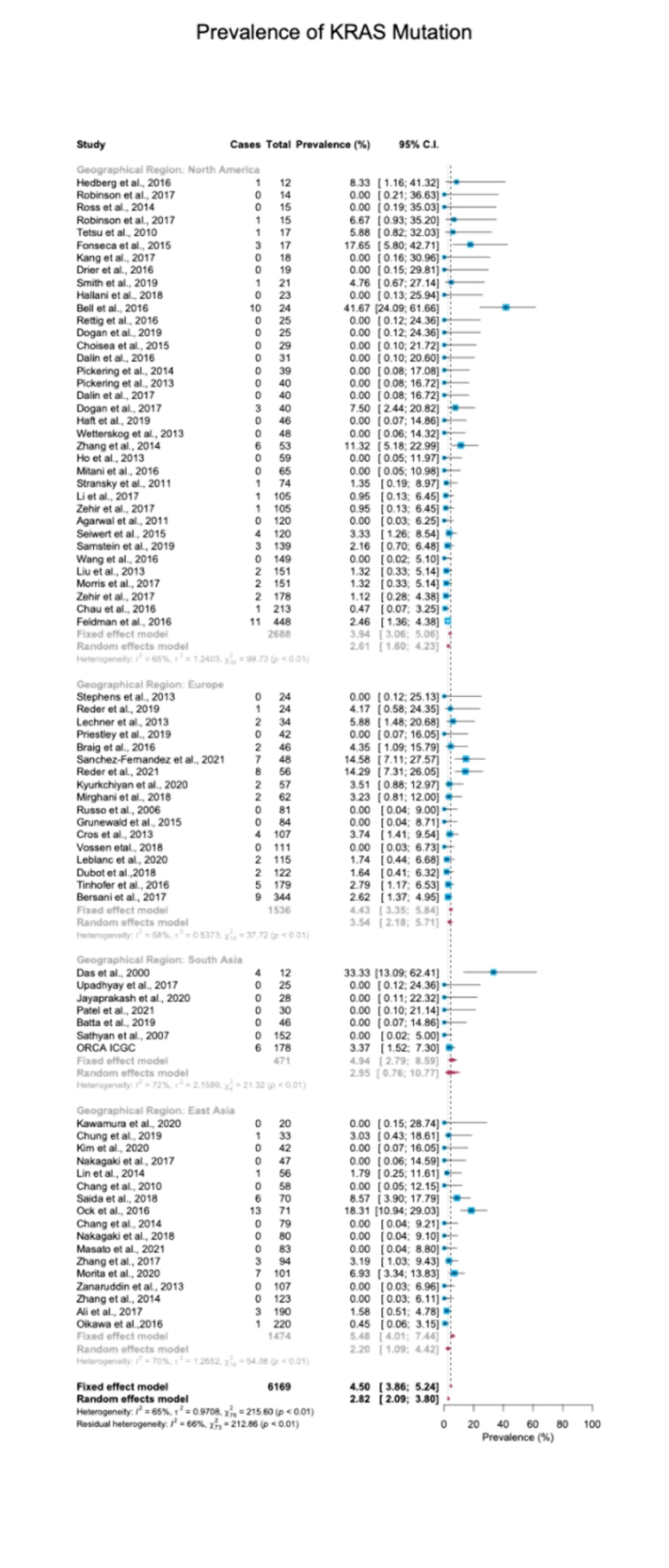

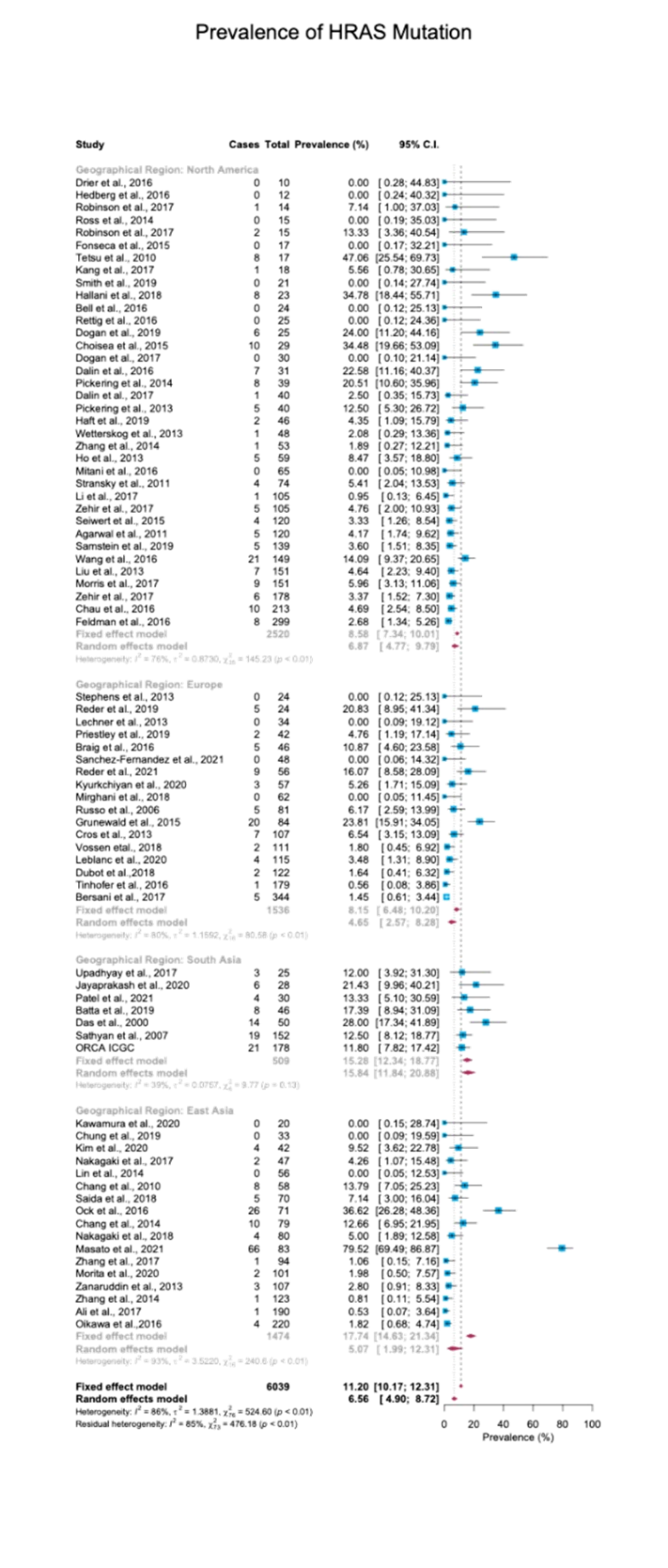

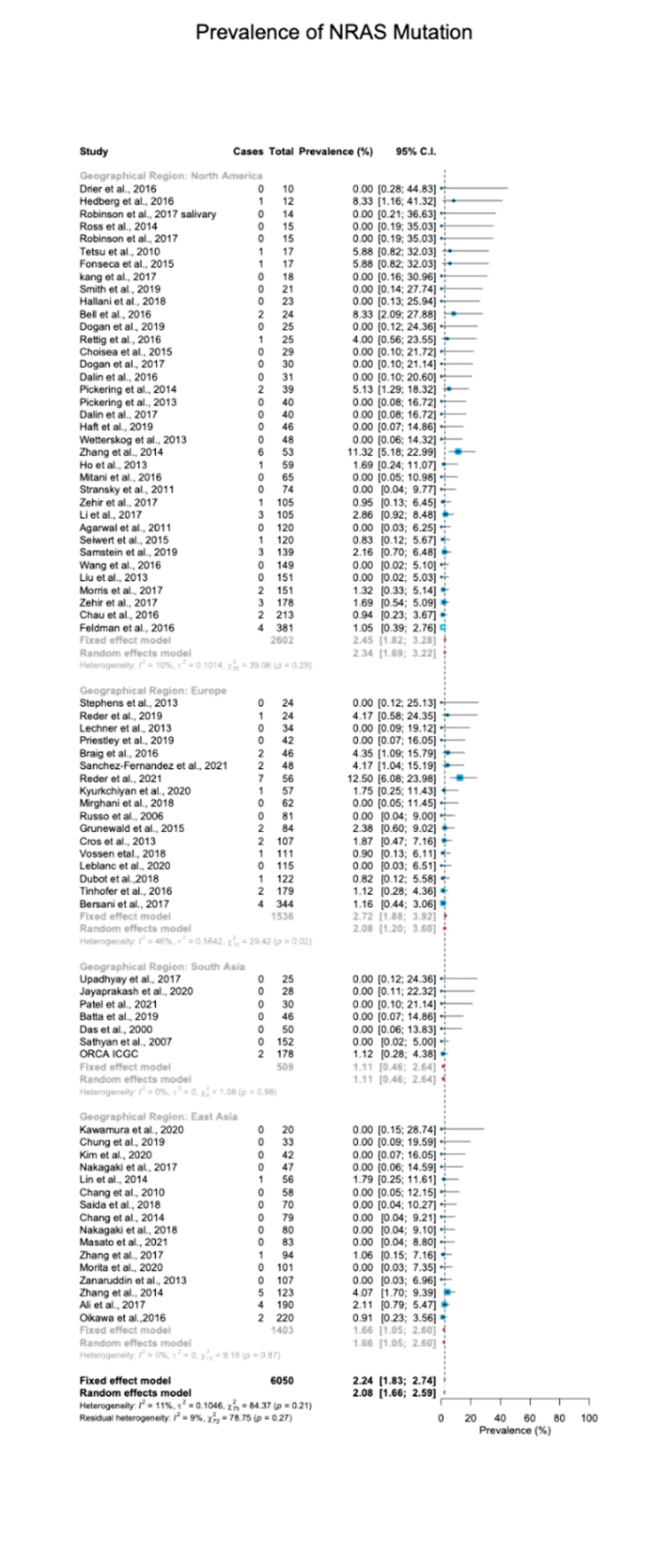
**HRAS	KRAS	NRAS**

**eFigure 3: Mutation prevalence according to anatomical site**

Forest plot of RAS mutation frequency [%] in head and neck cancer according to tumor anatomical site of origin. CI: Confidence interval. I^2^: Inconsistency index.

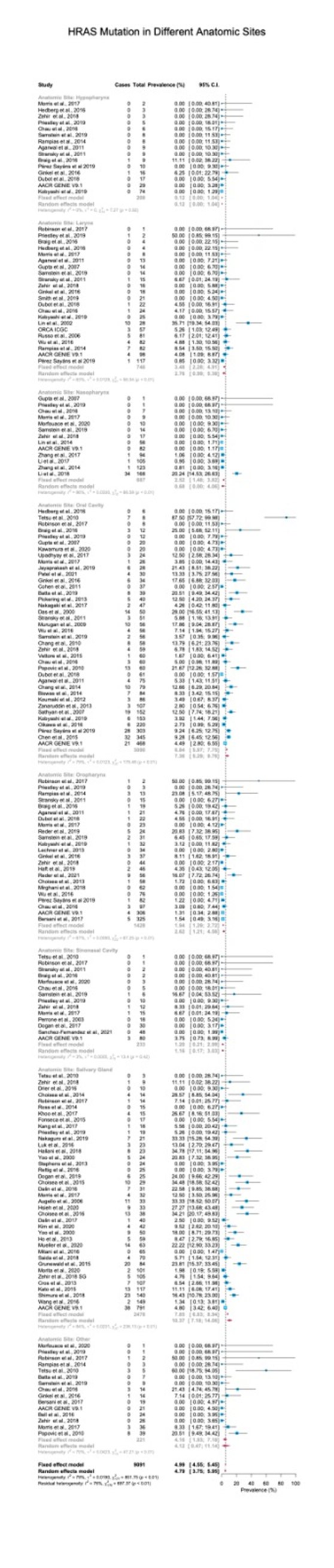

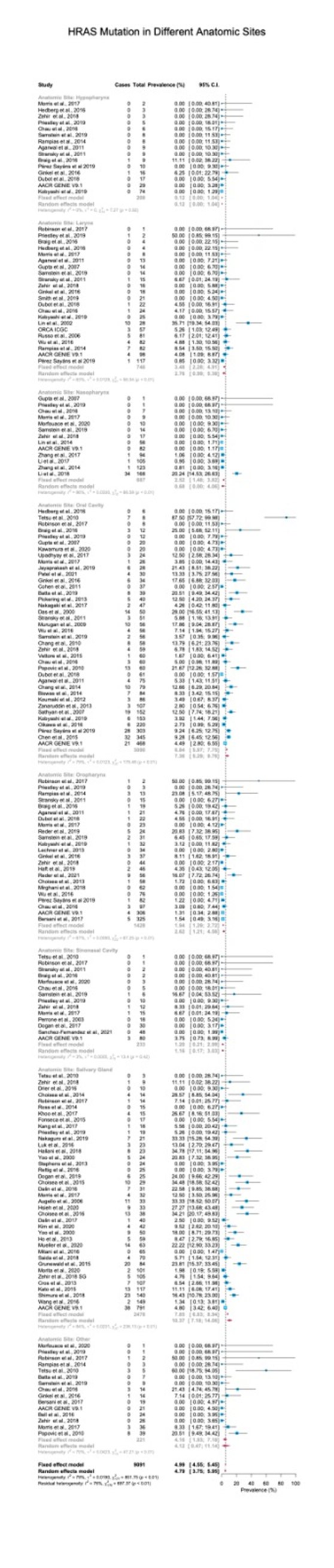

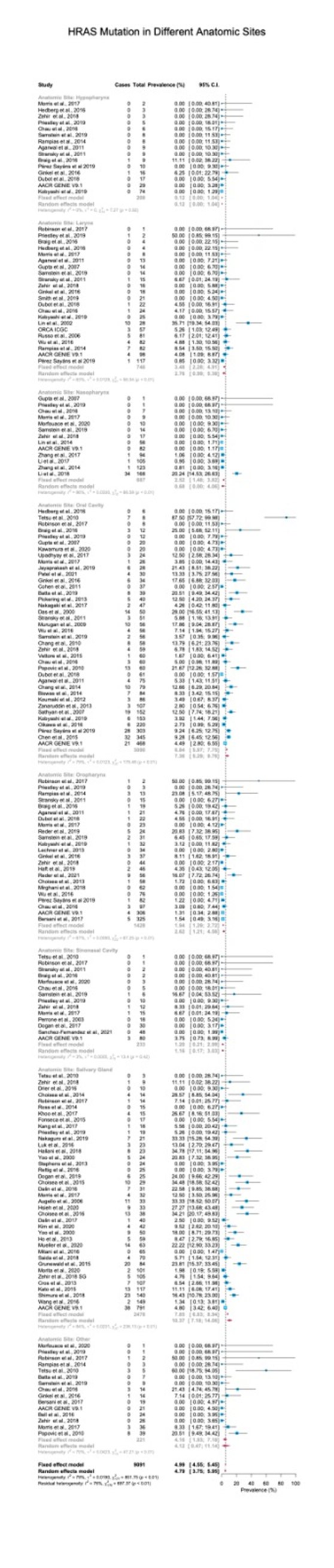
**HRAS**

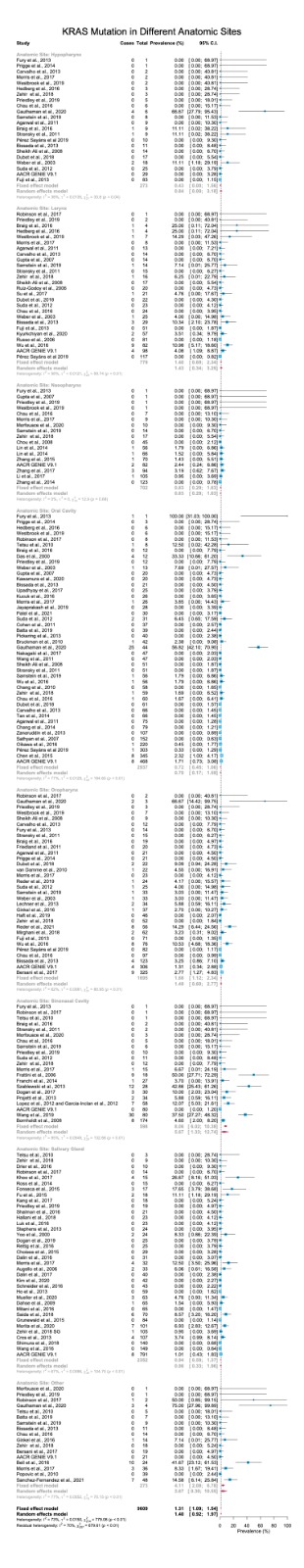

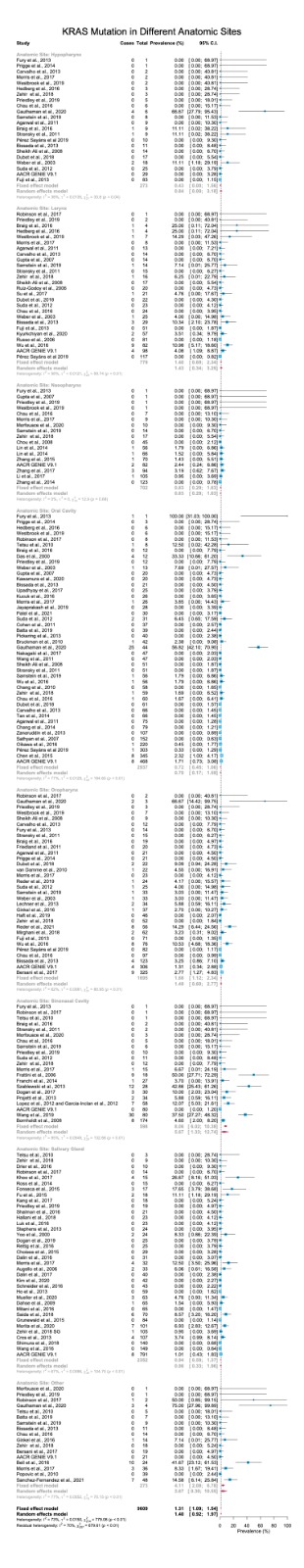
**KRAS**

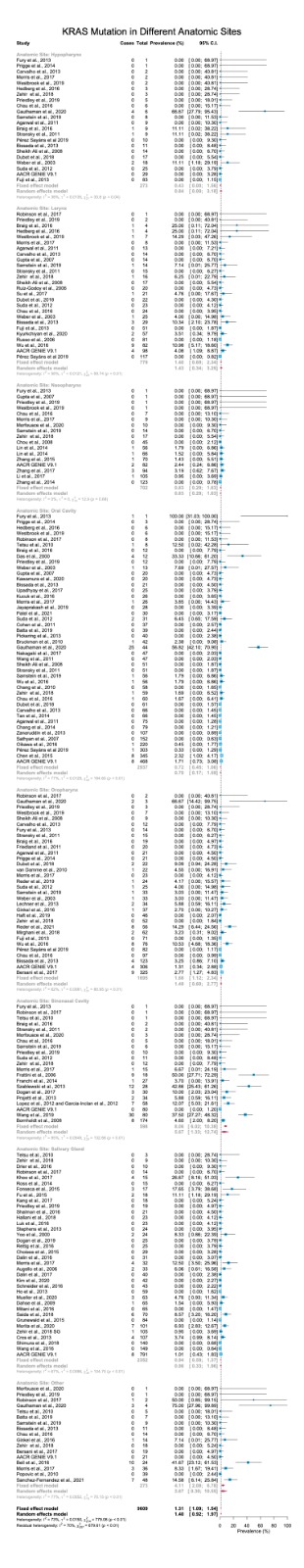

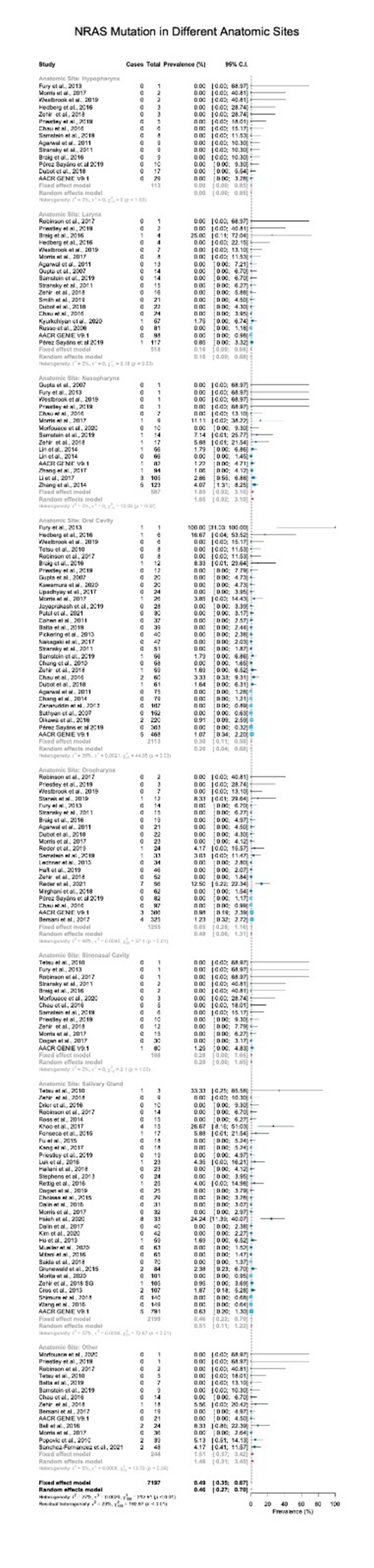
**NRAS**

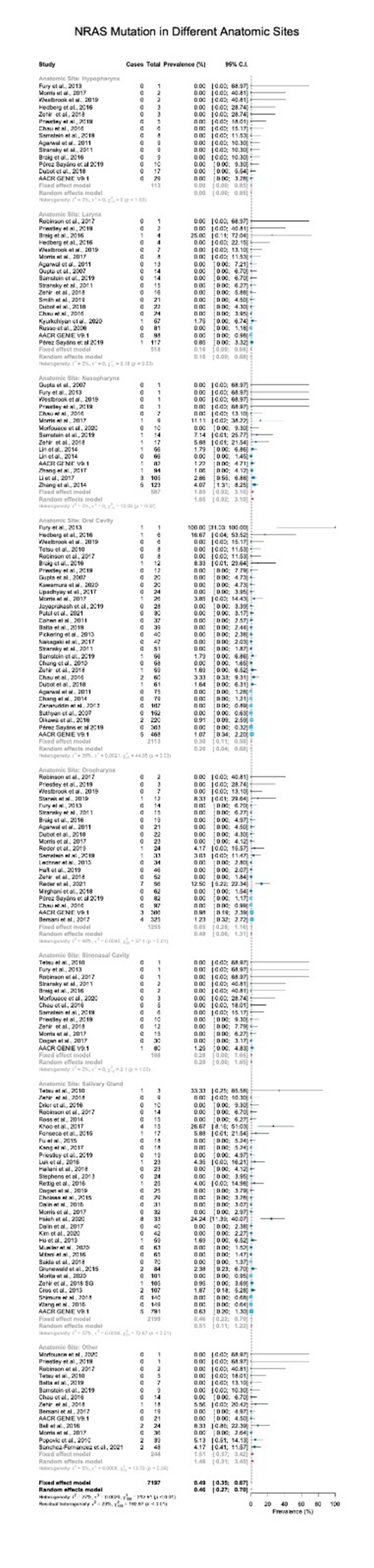

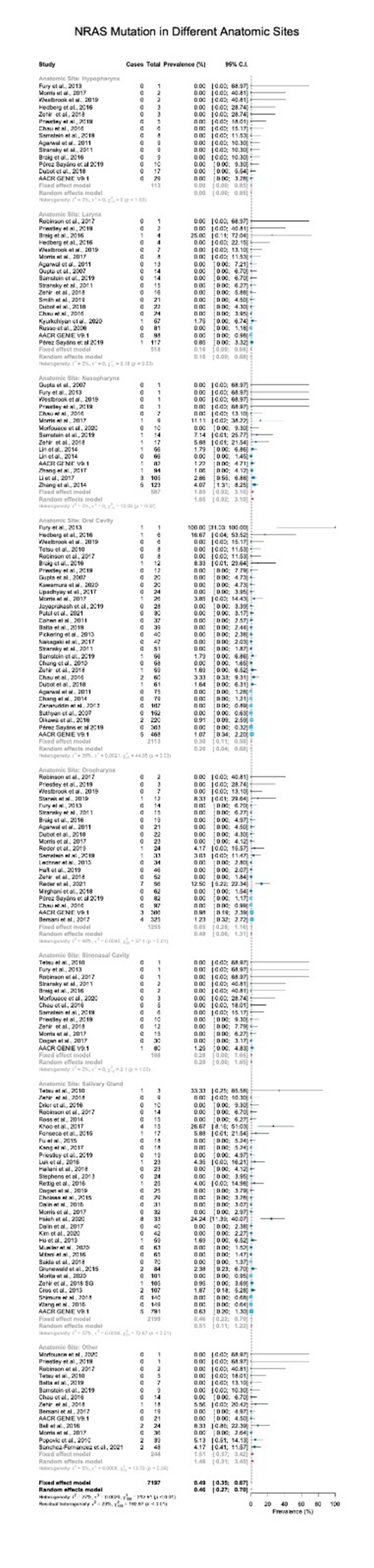

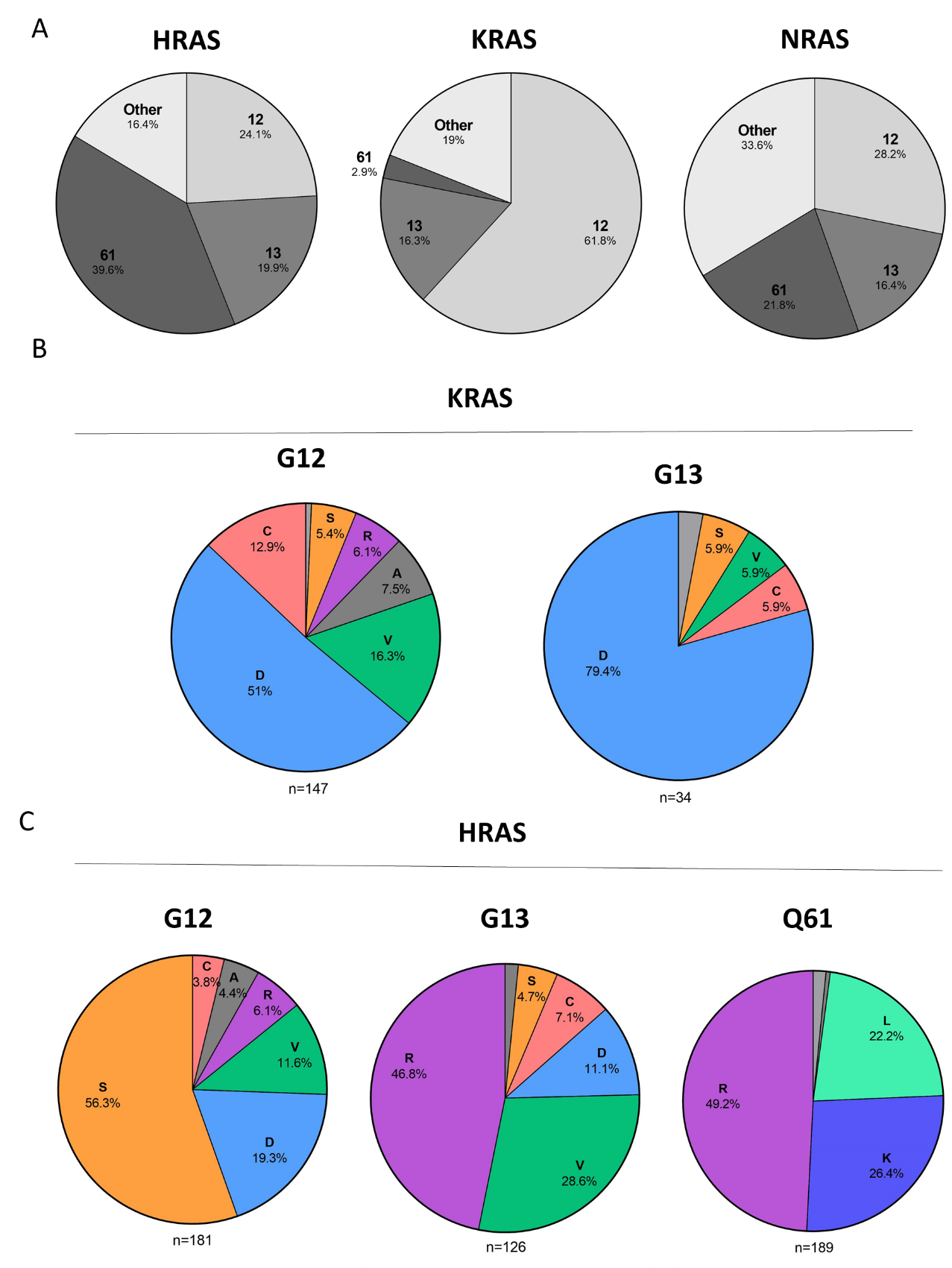
**eFigure 4: Amino Acid Substitutions**

**(A)** Mutated codons [%] in cases with HRAS, KRAS, and NRAS mutations.

**(B)** Amino acid substitutions [%] in cases with KRAS G12 and G13 mutations.

**(C)** Amino acid substitutions [%] in cases with HRAS G12, G13, and Q61 mutations.

D - aspartic acid, C - cysteine, V -valine, S - serine, R - arginine, A - alanine, K - lysine, L - leucine.

**eFigure 5: Association between RAS Mutations and Disease Stage/Grade**

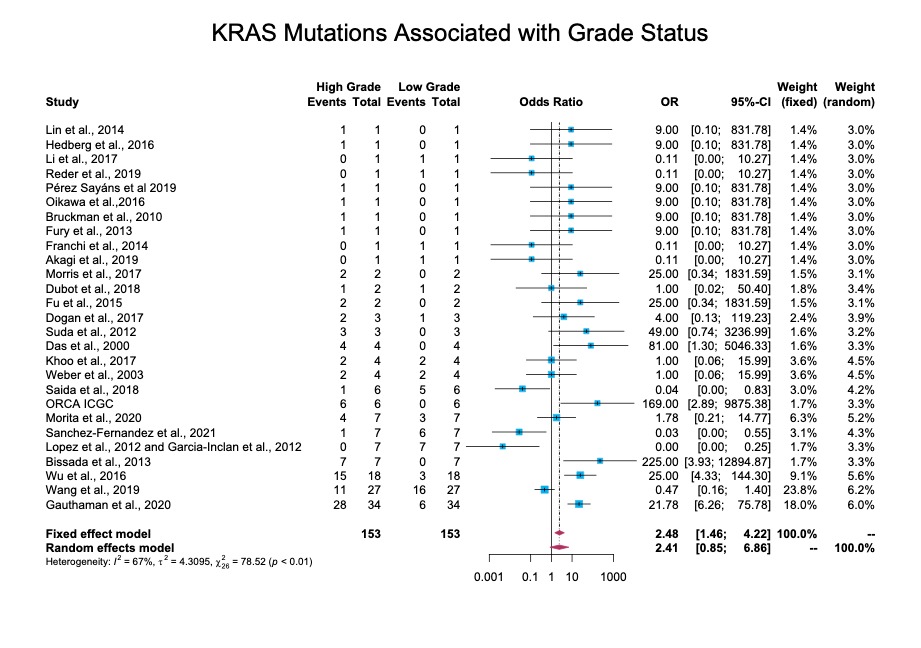
An odds ratio analysis of the association between tumor grade and KRAS or NRAS mutations; no statistically significant correlations were found.

**KRAS**

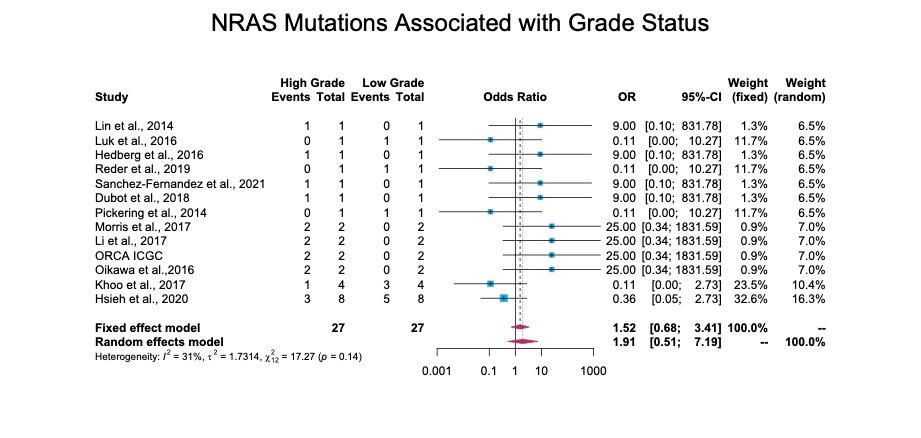
**NRAS**

**eFigure 6: Association between RAS Mutations and HPV status**

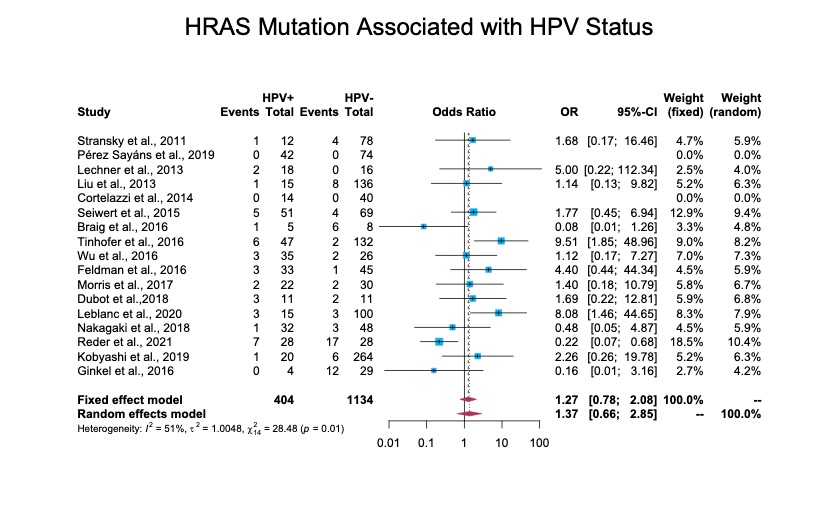
An odds ratio analysis of the association between human papillomavirus infection status and HRAS or NRAS mutations; no statistically significant correlations were found.

**HRAS**

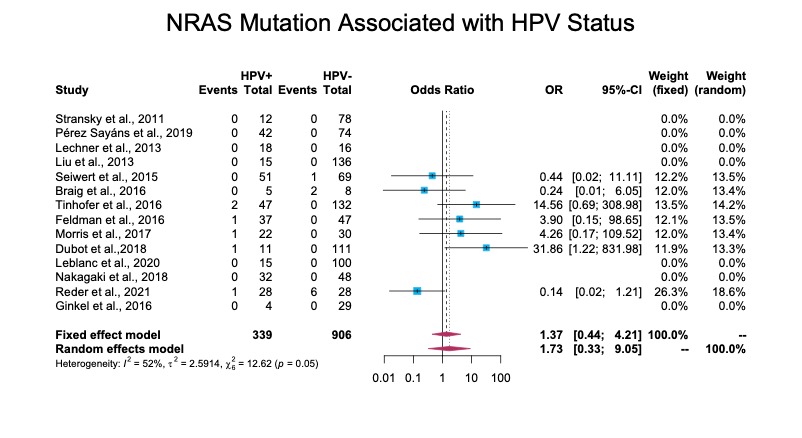
**NRAS**
